# A high content microscopy-based platform for detecting antibodies to the nucleocapsid, spike and membrane proteins of SARS-CoV-2

**DOI:** 10.1101/2021.10.14.21264873

**Authors:** Daniel M. Williams, Hailey Hornsby, Ola M. Shehata, Rebecca Brown, Domen Zafred, Amber S.M. Shun-Shion, Anthony J. Hodder, Deepa Bliss, Andrew Metcalfe, James R. Edgar, David E. Gordon, Jon R. Sayers, Martin J. Nicklin, Paul J. Collini, Steve Brown, Thushan I. de Silva, Andrew A. Peden

**Affiliations:** School of Bioscience, University of Sheffield, Western Bank, Sheffield, S10 2TN, United Kingdom; Department of Infection, Immunity and Cardiovascular Diseases, University of Sheffield Medical School, Beech Hill Road, Sheffield, S10 2RX, United Kingdom; Department of Pathology, University of Cambridge, Cambridge, CB2 1QP, United Kingdom; Department of Pathology, Emory University, Whitehead Building, Atlanta, GA, USA; South Yorkshire Regional Department of Infection and Tropical Medicine, Sheffield Teaching Hospitals NHS Foundation Trust, Glossop Road, Sheffield, S10 2JF, United Kingdom

## Abstract

The strong humoral immune response produced against the SARS-CoV-2 nucleocapsid (N) and spike (S) proteins has underpinned serological testing but the prevalence of antibody responses to other SARS-CoV-2 proteins, which may be of use as further serological markers, is still unclear. Cell-based serological screening platforms can fulfil a crucial niche in the identification of antibodies which recognise more complex folded epitopes or those incorporating post-translation modifications which may be undetectable by other methods used to investigate the antigenicity of the SARS-CoV-2 proteome. Here, we employed automated high content immunofluorescence microscopy (AHCIM) to assess the viability of such an approach as a method capable of assaying humoral immune responses against full length SARS-CoV-2 proteins in their native cellular state. We first demonstrate that AHCIM provides high sensitivity and specificity in the detection of SARS-CoV-2 N and S IgG. Assessing the prevalence of antibody responses to the SARS-CoV-2 structural membrane protein (M), we further find that 85% of COVID-19 patients within our sample set developed detectable M IgG responses (M sensitivity 85%, N sensitivity 93%, combined N + M sensitivity 95%). The identification of M as a serological marker of high prevalence may be of value in detecting additional COVID-19 cases during the era of mass SARS-CoV-2 vaccinations, where serological screening for SARS CoV-2 infections in vaccinated individuals is dependent on detection of antibodies against N. These findings highlight the advantages of using cell-based systems as serological screening platforms and raise the possibility of using M as a widespread serological marker alongside N and S.

## Introduction

Severe acute respiratory syndrome coronavirus 2 (SARS-CoV-2), the causative agent of Coronavirus Disease 2019 (COVID-19), has impacted the lives of nearly everyone in the developed world (Wu, Zhao et al. 2020). Controlling SARS-CoV-2 infections within communities and across populations requires accurate knowledge of both current and previous cases of COVID-19. Serological assays fulfil critical roles towards the latter, identifying individuals previously exposed to the virus who may potentially be immune whilst also contributing towards the construction of accurate epidemiological estimates of infection rates. Analysis of COVID-19 patient serum obtained through serological sampling has also provided invaluable information on the kinetics and profile of antibody responses produced against SARS-CoV-2 in relation to disease outcomes (Long, Liu et al. 2020, Long, Tang et al. 2020, Ripperger, Uhrlaub et al. 2020). Antibody tests of high sensitivity and specificity capable of providing a comprehensive overview of the humoral immune landscape induced after exposure to SARS-CoV-2 are therefore desirable to track the spread of the virus within communities and gain deeper insight into the interplay between virus and host immune system during acute and convalescent phases of infection.

To date, SARS-CoV-2 population-level serological testing relies upon detection of antibodies against either the SARS-CoV-2 nucleocapsid (N) or spike (S) proteins as both induce detectable humoral immune responses in the vast majority of those infected (National 2020). The serological assays currently in use for SARS-CoV-2 display varying levels of sensitivity and specificity and come with a variety of strengths and weaknesses (Krammer and Simon 2020). Screening for reactivity of patient sera against purified versions of the SARS-CoV-2 N and S proteins by ELISA is considered the gold standard for identification of patients who have previously been infected with SARS-CoV-2 (Amanat, Stadlbauer et al. 2020). However, this technique is limited in the scope of viral antigens which can be assayed due to the difficulty of producing viral proteins with physiochemical properties compatible with their production in a purified form (Johari, Jaffe et al. 2021). Although not employed for mass serological testing, additional techniques such as viral peptide and protein arrays have been helpful in identifying other immunogenic SARS-CoV-2 proteins. In contrast to ELISA based methods, viral peptide and protein arrays can provide almost complete coverage of the viral proteome allowing a more global overview of the humoral immune response and fine mapping of antibody epitopes, the latter being of particular of importance for looking at antibody cross-reactivity with other human coronaviruses and understanding the mechanism of action of neutralizing antibodies (Jiang, Li et al. 2020, Shrock, Fujimura et al. 2020, Ladner, Henson et al. 2021, Li, Xu et al. 2021, Stoddard, Galloway et al. 2021). The use of peptide fragments as bait however, limits detection of antibodies to those dependent on binding to contiguous stretches of amino acids. An additional drawback is that the proteins and peptide fragments used in both ELISA and protein or peptide arrays often do not include post translational modifications which may be important features of viral epitopes recognised by host antibodies.

Cell based methods such as immunofluorescence microscopy or flow cytometry can overcome some of the limitations associated with ELISA and viral peptide arrays and as such have been applied to different degrees to screen for the presence of antibodies induced after pathogenic infection (Meyer, Drosten et al. 2014, Grzelak, Temmam et al. 2020, Hambach, Stahler et al. 2021). In contrast to ELISA and protein or peptide arrays, viral proteins expressed in mammalian cells are produced in their three dimensional fully folded state and are likely subject to the same post translational modifications viral proteins undergo in infected cells. Screening of serum samples by immunofluorescence microscopy has demonstrated high sensitivity and specificity when applied previously during the SARS-CoV and MERS outbreaks (Chan, Ng et al. 2004, Meyer, Drosten et al. 2014), however this methodology has to date largely been used as an adjunct to those techniques described above during the current SARS-CoV-2 pandemic (Grzelak, Temmam et al. 2020, Wolfel, Corman et al. 2020).

In this paper, we describe the development of a cell-based expression platform combined with automated high-content immunofluorescence microscopy (AHCIM) to identify the presence of antibodies to specific SARS-CoV-2 proteins and test its performance in comparison to ELISA based COVID-19 serological testing. Calibration of our automated system using StrepTagged SARS-CoV-2 N and S proteins demonstrated that automated immunofluorescence-based antibody screening could detect N and S antibodies in sera collected from COVID-19 patients with high sensitivity and specificity. Most interestingly, further application of this system identified antibodies in a significant number of COVID-19 serum samples against the SARS-CoV-2 membrane (M) protein, opening up the possibility of using M as a third high prevalence serological marker alongside N and S. Together, our findings demonstrate the versatility and utility of a cell-based immunofluorescence assay for use as both a tool in seroprevalence studies and as an investigative tool to gain greater insight into humoral immune responses to pathogenic infection.

## Materials and Methods

### Antibodies and plasmids

For immunofluorescence microscopy experiments StrepTagged proteins were detected using StrepTactin-DY549 (1:1000, IBA Lifesciences 2-1565-050) and human IgG with a rabbit anti-human-IgG AlexaFluor-488 (1:500, Life technologies A1101). Primary antibodies used for western blotting were mouse anti-SARS-CoV-2 nucleocapsid (Genetex, GTX632269), mouse anti-strep-mAb (1:1000, IBA Lifesciences) and rabbit anti-GAPDH (Proteintech, 60004-1-Ig). Secondary antibodies for western blotting were HRP conjugated donkey anti-human IgG (1:2000, Biolegend 410902), HRP conjugated anti-mouse (1:2000, Jackson Immuno Research 115-035-008) and HRP-conjugated goat anti-rabbit (1:2000, Jackson Immuno Research 111-035-144). StrepTagged SARS-CoV-2 S, N and M expression constructs were a kind gift from Professor Nevan Krogan, University of California. All SARS-CoV-2 proteins were human codon optimised and expressed from pLVX-EF1alpha-IRES-puro (N and M) or pTwist-EF1alpha-IRES-puro (S) plasmids as previously described (Gordon, Jang et al. 2020).

### Recruitment and consent

Serum samples used were from healthcare workers (HCWs) recruited at Sheffield Teaching Hospitals NHS Foundation Trust (STH) as part of the COVID-19 Humoral Immune Responses in front-line HCWs (HERO) study, sampled in May and June 2020 (Hodgson, Colton et al. 2021). Regulatory approval was provided by HRA and Health and Care Research Wales (20/HRA/2180, IRAS ID: 283461). Anonymised serum samples from hospitalised COVID-19 patients and serum collected before 2017 during routine clinical care were used for assay validation purposes with approval from the STH R&D office as per standard practice.

### Cell culture and transfections

HEK-293T cells (CRL-3216) originally obtained from ATCC, were grown in Dulbeccos modified Eagles medium (Merck, D6429) supplemented with 10% fetal bovine serum (Gibco, 16140071), 100IU / ml penicillin, 100 μg/ ml streptomycin and 2mM glutamine (Merck, G1146-100ML). Cells were cultured at 37°C in a 5% CO_2_ humidified incubator.

### Immunofluorescence microscopy

For initial pilot experiments to assess the ability of immunofluorescence microscopy to detect SARS-CoV-2 antibodies in patient sera, cells were seeded onto poly-l-lysine (Sigma, P8920) coated glass coverslips and left to adhere overnight at 37°C before transfection. The next day, plasmid DNA was mixed with transfection reagent (Fugene HD, E2311) at a ratio of 1 μg DNA : 3ul Fugene in Optimem (GIbco, 31985-070) and incubated at room temperature for 10 minutes. The DNA-Fugene mixture was then diluted 1:10 in complete DMEM (10% fetal bovine serum (FBS), 1% penstrep) and added to cells. Cells were left at 37°C for a minimum of 16 hours before being processed for immunofluorescence microscopy.

Following transfection, the cell culture media was aspirated, and the cells and coverslips washed twice with phosphate buffered saline (PBS) to remove residual media. Cells were then fixed for 15 minutes at room temperature with 4% paraformaldehyde (PFA; Park Scientific Limited, 04018-4) in PBS, PFA quenched with 100mM glycine (Fisher Scientific, 10070150) for 5 minutes and cells permeabilised and blocked with a 0.1% Saponin (Sigma, S4521) and 1% bovine serum albumin (BSA) (Fisher Scientific, BP1600-100) solution (diluted in PBS) (IF buffer) for 10 minutes. For detection of intracellular proteins (N and M), coverslips were incubated with patient serum diluted at 1:125 in IF buffer for 1 hour at room temperature. For detection of cell surface S, coverslips were incubated with patient serum (1:125 in DMEM) for 1 hour at 37 °C prior to fixation. The patient serum was aspirated, and the coverslips were washed three times over 15 minutes using IF buffer. Bound antibodies and StrepTagged viral proteins were detected by incubating the coverslips with an anti-human IgG secondary antibody conjugated to AlexaFluor-488 (1:500, Life technologies A11014) and StrepTactin-DY549 (1:1000, IBA lifesciences 2-1565-050, 1 in 500 in IF buffer) for 1 hour at room temperature. Cell nuclei were then counterstained with DAPI (Sigma, D9542), coverslips washed a further 3 times for 5 minutes each with IF buffer and mounted on microscope slides with Prolong Gold Antifade Mountant (ThermoFisher scientific, P36930). Patient samples were imaged using a 20x objective (Olympus BX61 motorised wide-field epifluorescence microscope) and images collected with a Hamamatsu Orca monochrome camera.

### Manual quantification of immunofluorescence images

Non-transfected cells served as internal specificity controls for each patient serum incubation and were used to calculate background non-specific antibody staining. For manual quantification of signal strength from patient antibodies recognising SARS-CoV-2 N and S proteins, average 488-fluorescence intensity measurements were obtained in FIJI-ImageJ (Schindelin, Arganda-Carreras et al. 2012) from a minimum of six non-transfected and six transfected cells by drawing around the perimeter of each cell of interest and measuring the average 488-fluorescence intensity. Background fluorescence was then accounted for as follows. 488-fluorescence intensity measurements from the six non transfected cells per patient were averaged, and this value was then subtracted from each of the six 488-intensity measurements obtained from transfected cells measurements per patient. A ratio of cell associated fluorescence from non-transfected cells compared to transfected cells was obtained by dividing 488-intensity measurements from transfected cells identified by StrepTactin 549 staining by 488-intensity measurements from non-transfected cells. For manual quantification of antibodies recognising M, 488-signal associated with Golgi localised M identified by the StrepTag staining was quantified from a minimum of six transfected cells. For non-transfected cells 488-signal from a small perinuclear region of equivalent size was measured and a ratio calculated as described above for N and S.

Of note, our initial experiments assaying IgG responses against N suggested patient sera N antibody levels were limiting as we observed that having too many transfected cells significantly reduced the number of virus specific antibodies bound to transfected cells, as indicated by a weaker anti-human IgG-1 Alexa-488 signal at higher transfected cell densities (**Supplementary Figure 1**). To avoid using larger volumes of patient sera to boost the transfected cell specific Alexa-488 signal, in all other experiments using tagged SARS-CoV-2 proteins we instead controlled the ratio of transfected to non-transfected cells so that there was a significant excess of non-transfected cells (ratio of 10:1 non transfected: transfected cells).

### Automated high content microscopy and image analysis

HEK cells seeded in 6 well plates were transfected with SARS-CoV-2 plasmid DNA encoding either S, N or M proteins or mock transfected as described above, Cells were detached by rinsing with conditioned medium 24 hours after transfection. Mock transfected cells were mixed with SARS-CoV-2 protein transfected cells at a ratio of 10:1 before being seeded into a poly-l-lysine coated 96 well plate (Greiner-bio-one, 655098) at a density of 30,000 – 40,000 cells per well. Cells were then left to adhere overnight at 37°C

The next day, cells were processed for immunofluorescence microscopy as described above with a maximum of 96 serum samples analysed per plate. Following labelling of bound human IgG and StrepTagged SARS-CoV-2 proteins, images were acquired with a 20x objective on a Molecular Devices Image Xpress Micro high content microscope, collecting signal from DAPI, FITC and Texas red channels. For image acquisition, wells to be imaged were selected within the microscope software and a minimum of 4 sites per well imaged using a 20x objective. Captured images were analysed using the Molecular Devices Image Xpress Micro software package. Transfected and non-transfected cells were identified as described in Figure 2. Total fluorescence signal for each cell identified by the software was divided by cell area and an average fluorescence intensity for transfected and non-transfected cells calculated from multiple cells per serum sample assayed.

### Western blotting

HEK-293T cells expressing the nucleocapsid protein grown in 6 or 12 well dishes were detached by rinsing with conditioned media and cell suspensions collected in 15ml falcon tubes. Cells were then pelleted by centrifugation at 1000 *x g* for 10 minutes and the conditioned media aspirated from the pellet. Following one wash with PBS, the cell pellet was lysed in SDS sample buffer containing 5% β-mercaptoethanol and boiled for 10 minutes at 95°C. Cell lysates were then loaded onto a Tris-Glycine polyacrylamide gel and proteins resolved by SDS-PAGE gel electrophoresis. Gels were transferred overnight via wet transfer in 1xTobin buffer (25 mM Tris, 192 mM glycine, 20% Methanol) onto PVDF membranes (Cytiva, 10600023) before blocking in PBS containing 0.1% Tween20 (Sigma, P7949) (PBST) and 5% skim powder milk (Sigma, 70166) for 1 hour at room temperature. Membranes were then probed with either patient sera diluted 1:1000 in blocking solution or an anti-StrepTag monoclonal antibody (1:1000, IBA lifesciences 2-1507-001) overnight or for 1 hour at room temperature. Membranes were washed three times for 5 minutes each with PBST and incubated with either donkey anti-human (1:2000, Biolegend 410902) or anti-mouse (1:2000, Jackson Immuno Research 115-035-008) HRP conjugated secondary antibodies for a further 1 hour. After three 5 minute washes with PBST, membranes were incubated with Clarity Western ECL substrate (Bio-rad, 170-5061) and signal detected using a LICOR c-DiGiT imaging system. Quantification of western blots was performed using the Image Studio Lite software package.

### ELISA

ELISA for S and N proteins were performed as previously described (Hodgson, Colton et al. 2021). High binding microtiter plates (either Immulon 4HBX; Thermo Scientific, 6405, or Nunc MaxiSorp; Thermo Scientific, 442404) were coated overnight at 4oC with 50 μl/well SARS-CoV-2 protein diluted in PBS (pH 7.4). Either full-length spike produced in mammalian cells (Johari, Jaffe et al. 2021), or nucleocapsid produced in *E*.*coli* were used as antigens. Recombinant nucleocapsid protein was produced as previously described (Hodgson, Colton et al. 2021). Once coated, plates were washed 3x with 0.05% PBS-Tween, and blocked for 1 hour with 200 μl/well casein blocking buffer. Plates were emptied (no wash step) and loaded with 100 μl/well of samples and controls, diluted to 1:200 for a 2 hour incubation. Plates were washed 3x and loaded with 100 μl/well goat anti-human IgG-HRP conjugate (Invitrogen, 11594230) at 1:500 dilution for a 1 hour incubation. After a final 3x wash, plates were developed with 100 μl/well TMB substrate (KPL, 5120-0074) for 10 minutes in the dark. The reaction was stopped by addition of 100 μl/well 1% HCl stop solution (KPL, 5150-0021). Absorbances were read immediately at 450 nm.

### Statistical analysis

All graphs included in figures were made using the GraphPad Prism software package. Statistical comparisons between groups using an unpaired t-test and pearsons correlation coefficient analysis of N, S and M antibody relationships were all performed using GraphPad Prism.

## Results

### Detection of SARS-CoV-2 antibodies using immunofluorescence microscopy

A recent complication in SARS-CoV-2 serological testing has arisen from the use of S as the immunogen applied in SARS-CoV-2 vaccination programs. This prevents the use of S as a marker of past infection and makes detection of prior SARS-CoV-2 infection in those vaccinated dependent on assaying for antibodies against N. The identification of additional SARS-CoV-2 serological markers of high seroprevalence would be beneficial under these circumstances to ensure robust high sensitivity serological testing. Due to limitations involved in the use of other methods applied to characterise SARS-CoV-2 humoral immune responses, the seroprevalence and strength of antibody responses against other SARS-CoV-2 proteins is unclear with no other serological markers of high prevalence having been identified to date (Jiang, Li et al. 2020, Shrock, Fujimura et al. 2020). In addition to being of use as an additional method to screen for the presence of antibodies to N and S, cell-based expression platforms are an ideal system in which to test the antigenicity of other SARS-CoV-2 proteins; SARS-CoV-2 proteins transiently expressed in mammalian cells (Gordon, Hiatt et al. 2020, Gordon, Jang et al. 2020) can be assayed for reactivity with patient sera in a state likely to closely resemble that encountered by the humoral immune system. Such a system also bypasses the requirement for purified antigens. Given its relative simplicity, we investigated the utility of an immunofluorescence microscopy-based method to detect antibodies against specific SARS-CoV-2 proteins in human serum samples (**Figure 1A and B**). To test the effectiveness of this approach, we transfected HEK-293 cells with codon optimised plasmids encoding StrepTagged SARS-CoV-2 N and S (**Figure 1C and F)**. Based on reports from the previous SARS-CoV pandemic of antibody responses directed towards the SARS-CoV M protein (Wang, Wen et al. 2003, He, Zhou et al. 2005), we also investigated whether COVID-19 patients develop IgG responses against SARS-CoV-2 M (**Figure 1I**). Cells transfected with either StrepTagged N or M were fixed, permeabilised and incubated with sera from either pre pandemic individuals or from samples confirmed to have been infected with SARS-CoV-2 by RT-PCR. Cells transfected with a StrepTagged S protein were incubated with sera from the same groups prior to fixation. Transfected cells were identified using StrepTactin labelled with DY-549 and the antigen specific antibodies using anti-human IgG1 secondary antibody conjugated to Alexa488.

**Figure 1.**
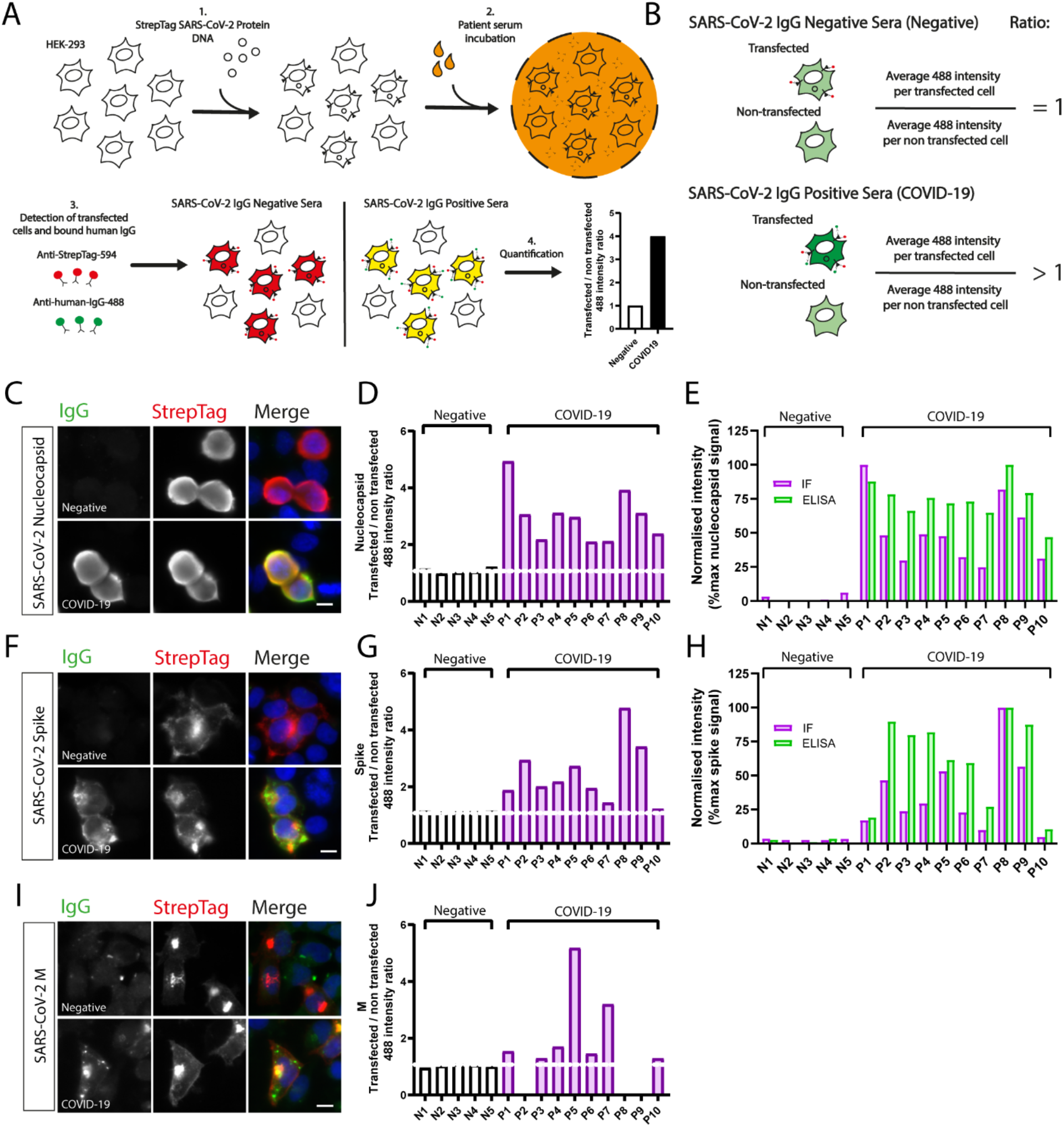
Detection of antibodies against SARS-CoV-2 proteins using immunofluorescence microscopy. **(A)** Schematic of the work flow used to screen patient sera for SARS-CoV-2 antibodies by immunofluorescence microscopy. **(B)** Overview of how fluorescence intensity ratios used to estimate the relative strength of IgG responses against the indicated SARS-CoV-2 protein were calculated. Representative immunofluorescence microscopy images of HEK-293 cells transiently expressing StrepTagged SARS-CoV-2 nucleocapsid **(C)**, spike **(F)**, or M **(I)** incubated with either pre pandemic (Negative) or SARS-CoV-2 positive (COVID-19) patient sera. Bound human IgG and cells expressing StrepTag-Spike were detected with an Alexa488 labelled anti-human IgG secondary antibody and StrepTactin-549 respectively. Quantification of the ratio of Alexa488 labelled anti-human IgG signal intensity from StrepTag SARS-CoV-2 nucleocapsid **(D)**, SARS-CoV-2 spike **(G)** or SARS-CoV-2 M **(J)** transfected or non-transfected cells for 5 negative (N1-N5) and 10 COVID-19 (P1-P10) serum samples. Comparison of nucleocapsid **(E)** and spike **(H)** IgG responses measured by immunofluorescence with values generated by ELISA for the same samples. Due to limited amounts of sera, it was not possible to analyse all of the samples for M. Thus, some samples have been left blank in panel **(J)**. Scale bars=10μm.

As the transfection efficiency for the viral proteins was less than 100% we were able to use non-transfected cells as a control for non-specific signal (**Figure 1B**). As an indirect readout of relative antibody concentrations in the serum samples we assayed, we measured the fluorescence intensity of anti-human IgG-1 Alexa-488 signal associated with both transfected and non-transfected cells and calculated a ratio of transfected to non-transfected 488 fluorescence intensity. Pre-pandemic serum samples had ratios close to one, reflecting equal amounts of signal associated with transfected and non-transfected cells (non-specific background signal). As indicated by ratios of greater than one, antibodies specific to the N and S proteins were only observed in sera from individuals previously infected with SARS-CoV-2 (**Figure 1D and G**). Interestingly, we could also see that many serum samples from COVID-19 patients contained detectable IgG against the SARS-CoV-2 M protein (**Figure 1J**). To see how the results from our microscopy-based assay for N and S compared with those obtained using ELISA for the same set of samples, we normalised the data generated from both methods by expressing values as a percentage of the maximum signal within each dataset. Similar trends could be seen between ELISA absorbance readings and immunofluorescence ratios (**Figure 1E and H**). Quantitative immunoblotting of N signal in the same serum samples also closely matched the results observed by ELISA and IF (**Supplementary Figure 2A-C**). These preliminary results confirm that the combined use of transiently expressed tagged SARS-CoV-2 proteins with immunofluorescence microscopy is a viable approach to detect antibodies against SARS-CoV-2 antigens without the need for purified viral proteins.

### Developing an automated pipeline for quantifying the levels of antibodies to SARS-CoV-2 N, S and M proteins

Whilst screening serum samples by manual imaging and quantification provided proof of principle that immunofluorescence microscopy could be used to detect antibodies to viral antigens, the amount of work required to image and analyse samples manually would make it difficult to efficiently screen large sample numbers. To overcome this, we used a high content microscope in conjunction with automated image analysis software (automated high content immunofluorescence microscopy; AHCIM) (**Figure 2A**). Cells were identified in each captured image using a mask based on DAPI staining of the nuclei (**Figure 2B**). From the nuclear mask a series of additional masks were generated that were used to: 1) determine whether the cells were transfected based on a threshold value of StrepTactin549 signal and 2) measure the amount bound antibody. Applying these masks, the software was able to analyse hundreds of transfected and non-transfected cells per sample (**Figure 2B, C and D**). The average intensity for the transfected and non-transfected cells was generated and the ratio calculated for each sample. To validate the image analysis pipeline, we repeated the experiments shown in **Figure 1** using AHCIM and compared the data to the manual quantification. Ratios calculated automatically from images captured by AHCIM were comparable to those generated through manual quantification of the same serum samples (**Figure 2E, F and G**). These results demonstrate that the automated image capture and analysis workflow can accurately identify and quantify bound human IgG in COVID-19 patient sera. This methodology also significantly increases the throughput of immunofluorescence-based detection of SARS-CoV-2 antibodies in patient serum samples.

**Figure 2.**
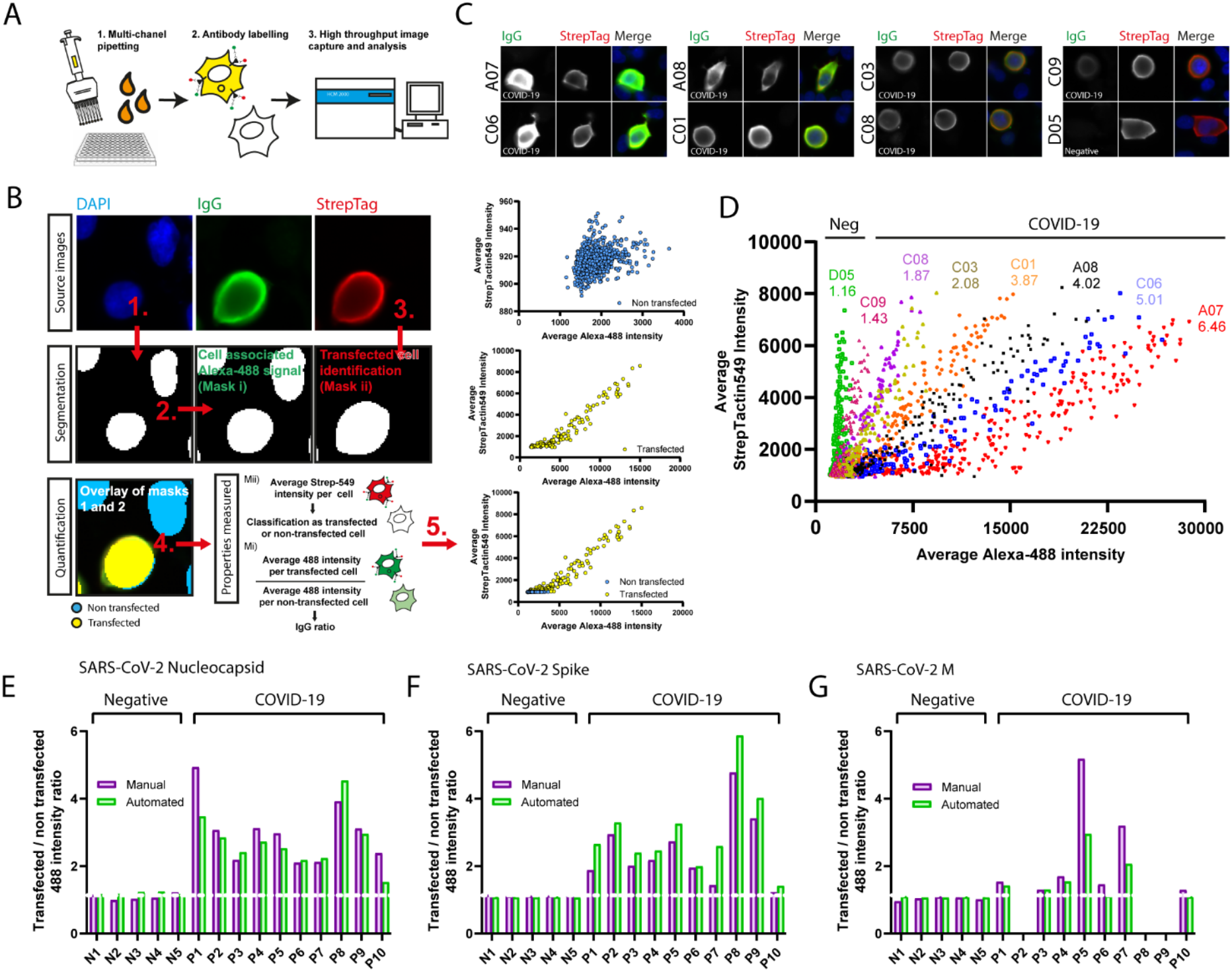
Application of high content microscopy and automated image analysis to IF based COVID-19 antibody screening. **(A)** Overview of workflow involved in processing samples for high content microscopy and automated image analysis and **(B)** summary of method used by automated image analysis software to segment cells and classify as either transfected or non-transfected. Representative images are from cells transfected with StrepTagged-Nucleocapsid and incubated with sera from a COVID-19 positive patient. Parameters measured by the software to calculate serum sample ratios are indicated after step 4. Representative scatter plots for transfected and non-transfected cells identified from a COVID-19 positive serum sample are shown after step 5. **(C)** Representative images of IgG signal associated with Nucleocapsid transfected cells following incubation with serum samples listed in **(D). (D)** Scatter plots of Alexa-488 versus StrepTactin549 signal measured for nucleocapsid transfected cells identified by automated image analysis incubated with either pre-pandemic sera (D05) and SARS-CoV-2 IgG positive sera (C09, C08, C03, C01, A08, C06, A07). Plots are representative of the number of cells analysed from one field of view captured using a 20X objective with cells at approximately 60-80% confluency. SARS-CoV-2 Nucleocapsid IgG positive serum samples range from weak positive to strong positive IgG responses. Comparison of 488 intensity ratios for **(E)** nucleocapsid, **(F)** spike and **(G)** M IgG levels in pre-pandemic and COVID-19 serum samples generated by manual and automated image quantification for training set 15 samples previously described in Figure 1.

### AHCIM displays high sensitivity and specificity in detection of SARS-CoV-2 N and S IgG

Having determined that AHCIM image capture and analysis could accurately quantify bound IgG against SARS-CoV-2 proteins, we applied this system to a larger sample set consisting of 258 samples containing serum from 62 individuals acquired prior to the SARS-CoV-2 pandemic (pre-pandemic negative controls) and 196 patients confirmed to have been infected by the SARS-CoV-2 virus by RT-PCR (serum samples taken 10 to 120 days from positive RT-PCR result). Using our automated platform, we observed a significant difference in signal intensity between the pre-pandemic negative samples and RT-PCR positive samples for N and S **(Figure 3A and E**). In both cases the positive signal was approximately three-fold over the observed background signal. To determine if the severity of the disease impacted the antibody titre we stratified the samples based on whether the patient was hospitalised or not. In comparison to COVID-19 outpatients, IgG levels were significantly elevated for both N and S in COVID-19 inpatient samples (**Figure 3B and Figure 3F**. Mean N inpatient IgG ratio; 4.98. P<0.0001 vs outpatient. Mean S inpatient IgG ratio; 3.86. P=0.003 vs outpatient). These trends were also seen in ELISA absorbances for the same inpatient or outpatient sample sets (**Supplementary figure 3A-3D**).

**Figure 3.**
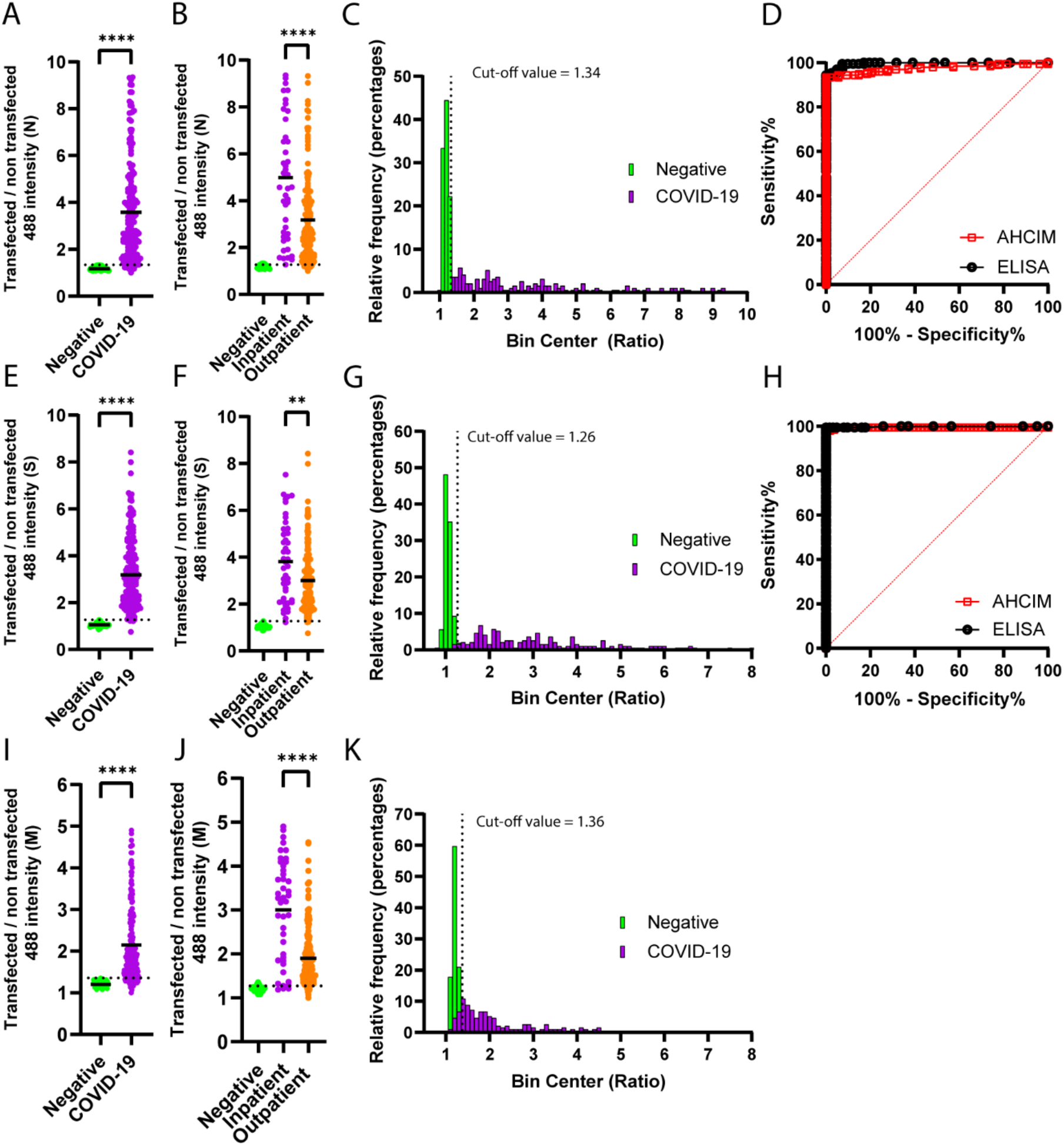
Immunofluorescence based high content microscopy and image analysis demonstrates high sensitivity and specificity for detection of SARS-CoV-2 Nucleocapsid, Spike and M IgG. To assess the performance of AHCIM to identify SARS-CoV-2 antibodies against nucleocapsid, spike and M across a larger set of serum samples, HEK293-T cells were transfected with the indicated StrepTagged SARS-CoV-2 constructs and incubated with patient sera from a sample set consisting of 62 COVID-19 negative pre-pandemic and 196 COVID-19 positive samples. Bound IgG was detected with an anti-human Alexa-Flour488 conjugated secondary antibody and imaged by automated high content immunofluorescence microscopy. Patient sample ratios were calculated as described in Figure 2 and plotted. Comparison of IgG ratios for negative and COVID-19 serum samples for **(A)** nucleocapsid **(E)** spike and **(I)** M. Stratification of COVID-19 samples based on inpatient (n=44) and outpatient (n=152) status and comparison of IgG ratios in inpatient and outpatient groups for **(B)** nucleocapsid **(F)** spike and **(I)** M. Dashed lines on Y axis indicate the calculated AHCIM cut-off values for each antigen. Frequency distribution histograms of calculated nucleocapsid **(C)**, spike **(G)** and M **(K)** IgG ratios from negative and COVID-19 positive patient serum samples. Ratios were sorted into bins of a width 0.1 ratio points for nucleocapsid, spike and M. Plots show the overlap between IgG ratios for pre-pandemic and COVID-19 positive samples. Detection of Spike IgG shows the smallest amount overlap between positive and negative IgG ratios, consistent with AHCIM based detection of S giving the highest specificity and sensitivity values. ROC curves comparing the sensitivity and specificity of IF and ELISA based detection of **(D)** nucleocapsid and **(H)** spike IgG in COVID-19 positive and pre-pandemic sera. \*\*\*\**P*<0.0001, \*\**P*<0.01 (P=0.0030) (Unpaired t-test).

Based on the IgG ratios calculated for each sample, we constructed receiver operating characteristic (ROC) curves for N and S to assess the overall sensitivity and specificity of our automated immunofluorescence-based platform in detecting N and S IgG (**Figure 3D and Figure 3H**). Inspection of the ROC curve generated for N indicated that a threshold of 1.34 provided the best trade-off between sensitivity and specificity for classification of samples as COVID19 positive or COVID19 negative. This threshold classified 182 out of 196 as COVID-19 positive based on the presence on detectable N IgG, misclassifying 14 samples as COVID-19 negative (**Supplementary Table 2)** (Sensitivity 92.9%, Specificity 100%. 95% CI; 88.4-95.7% sensitivity). AHCIM performed marginally better at reliably identifying positive and negative samples based on the presence of IgG to S (**Figure 3G & H**). Here, a cut-off value of 1.27 classified 192 out of 196 samples as COVID-19 positive (**Supplementary Table 2)** (Sensitivity 97.9%, Specificity 100%. 95% CI; 94.8-99.2% sensitivity). The better performance of the assay for S is most likely in part because assaying for S antibodies is performed on non-permeabilised cells which give lower levels of background fluorescence compared to permeabilised cells (**Supplementary figure 4A**).

To assess the performance of AHCIM in comparison to ELISA, we compared ROC curve AUC values from ELISA and AHCIM analysis of the same collection of 258 serum samples. ELISA based detection of N antibodies performed better than AHCIM at classifying samples as COVID19 positive or COVID19 negative (**Figure 3D**, N ELISA AUC = 0.99 vs N AHCIM AUC = 0.97). Optimal cut-off values prioritising high specificity for ELISA based analysis classified 185 samples as COVID-19 positive (Cut-off value = 0.72. 94.4% sensitivity, 100% specificity. 95% CI; 90.2-96.8% sensitivity), slightly higher than the 182 samples identified by AHCIM (**Supplementary Table 1 & 2**). ROC curve AUC values were similar between ELISA and AHCIM for COVID-19 inpatients, consistent with the higher N antibody levels observed in COVID-19 inpatients (**Figure 3B, Supplementary Figure 6A-B and Supplementary Tables 1 & 2;** ELISA N inpatient AUC = 1.00 vs AHCIM N inpatient AUC = 0.99). For detection of S antibodies, the performance of AHCIM based classification of samples as COVID-19 positive or negative based was comparable to ELISA (**Figure 3H and Supplementary Tables 1 & 2**; S ELISA AUC = 0.99 vs S AHCIM AUC = 0.99). 195 COVID-19 patient samples had above threshold S IgG levels when assayed by ELISA, 3 more than identified by AHCIM. Consistent with these results, minimal difference in S ROC curve AUC values were observed when samples were stratified based on inpatient or outpatient status (**Supplementary Figure 6C-D and Supplementary Tables 1 & 2;** ELISA inpatient AUC = 0.99 vs AHCIM inpatient AUC = 0.99. ELISA outpatient AUC 0.98 vs AHCIM outpatient AUC 0.99). As described for manual and automated quantification of the original training set 15 differences in antibody titres could also be reliably discerned within this larger collection of serum samples by AHCIM. There was high level of concordance between the strength of ELISA absorbances and AHCIM IgG ratios for both N and S in each serum sample tested (**Supplementary Figure 5A & B**). Collectively, these results demonstrate that AHCIM provides high specificity and sensitivity for detection of both SARS-CoV-2 N and S IgG.

### AHCIM reveals a significant proportion of COVID-19 patients have IgG against the SARS-CoV-2 M protein

As a high percentage of COVID-19 patients in our training set had antibodies against the SARS-CoV-2 M protein, we also tested our larger sample set to better establish the prevalence of antibody responses against SARS-CoV-2 M. As predicted from our pilot experiments we observed a significant difference in signal intensity between the pre-pandemic COVID-19 negative samples and RT-PCR positive samples for M (**Figure 3I**, pre-pandemic 1.20 +/- 0.06 S.D., COVID-19 positive 2.15 +/- 0.92 S.D. P<0.0001). Based on these results, we generated a ROC curve to determine the optimal cut-off value for classifying samples as COVID-19 positive or negative based on detection of M IgG (**Supplementary figure 6E**). A threshold of 1.36 provided 100% specificity and 84.7% sensitivity, classifying 166 out of 196 serum samples collected from COVID-19 patients as positive (M AHCIM AUC = 0.96, 95% CI; 79.0-89.1% sensitivity). A frequency distribution histogram of IgG ratios calculated for individual COVID-19 negative and positive samples revealed that 98.4% of ratios from negative samples fell within a range of 1-1.3 (**Figure 3K**). In contrast, ratios from 12.9% of COVID-19 positive samples fell between 1-1.3 with ratios greater than 1.3 accounting for the remaining 87.1% of COVID-19 positive samples. Similar to N and S, our automated platform was able to classify samples as COVID-19 negative or COVID-19 positive with greater confidence in COVID-19 inpatients (**Figure 3J, Supplementary Figure 6E, Supplementary Tables 1 & 2;** M inpatient mean ratio = 3.0 +/- 1.14 S.D., M outpatient mean ratio = 1.9 +/- 0.66 S.D. P<0.0001. M inpatient AHCIM AUC = 0.96, M outpatient AHCIM AUC = 0.96). As 166 samples from COVID-19 patients had ratios of greater than or equal to 1.36, these results indicate that across a larger population, 85% of SARS-CoV-2 infected individuals may develop IgG responses against SARS-CoV-2 M, much higher than could be previously inferred from other recent studies (Jiang, Li et al. 2020, Shrock, Fujimura et al. 2020).

### Combined use of N and M antibody testing boosts the sensitivity of AHCIM and ELISA based serological testing

Given the high prevalence of antibody responses to M observed by AHCIM, we examined whether the combined detection of antibodies to M and N could boost the sensitivity of COVID-19 serological testing in cases where S cannot be used to confirm prior SARS-CoV-2 infection. 159 of the 196 COVID-19 positive samples were positive for both N and M, less than the number detected using N or M individually, suggesting that the presence of detectable N IgG does not reliably predict the presence of detectable M IgG and vice versa. This was also evident from plots of N IgG ratios against M IgG ratios from COVID-19 serum samples. S and M antibody levels showed a closer correlation (R^2^=0.5201, P=<0.0001) than that between both N and M antibody levels (R^2^=0.3617, P=<0.0001) and N and S antibody levels (R^2^=0.3113, P=<0.0001) **(Figure 4A-J)**. Summing the ratios for N and M increased the sensitivity of AHCIM based antibody testing to 95.4%, detecting 187 out of 196 COVID-19 cases (Specificity 100%. 95% CI; 91.5-97.6% sensitivity), higher than the use the use of N or M individually (M sensitivity 84.7%, N sensitivity 92.9%) (**Figure 5C-D, Supplementary Table 3**). The combined detection of N and M antibodies therefore provides a boost to the sensitivity of AHCIM based antibody screening with these results suggesting that samples misclassified as negative by AHCIM based on N antibody levels can often be picked up screening for M antibodies and conversely, samples with a lack of detectable M antibodies can be picked up by screening for N.

**Figure 4.**
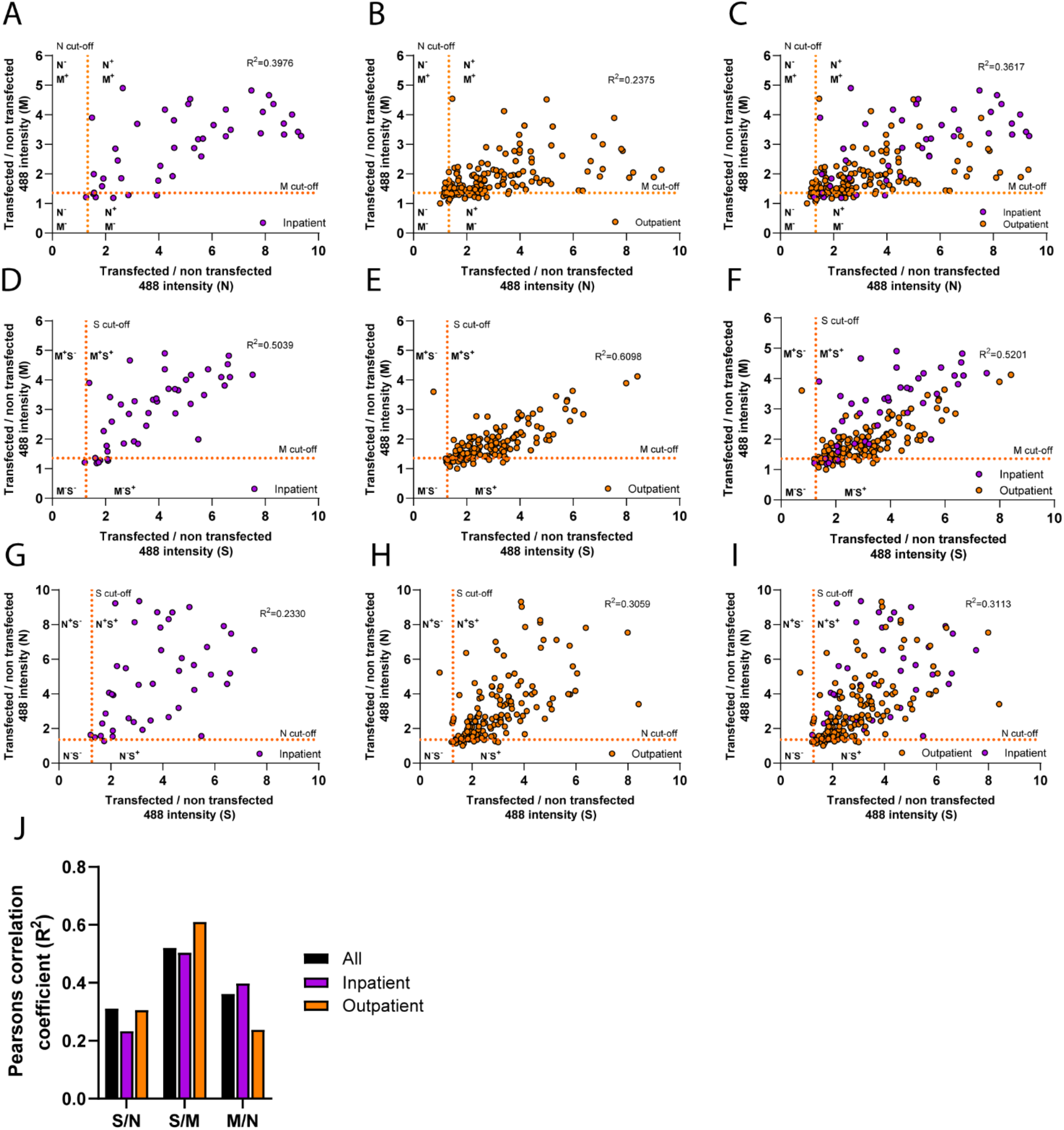
Relationship between N, M and S IgG levels. To examine the relationships between N, S and M antibody levels, IgG ratios from individual samples for N, S and M were plotted against one another in different combinations. M and N antibody levels per serum sample were plotted for **(A)** inpatients **(B)** outpatients and **(C)** all patients combined. S and M antibody levels per serum sample were plotted for **(D)** inpatients **(E)** outpatients and **(F)** all patients combined. S and N antibody levels per serum sample were plotted for **(G)** inpatients **(H)** outpatients and **(I)** all patients combined. Dashed orange lines on each graph indicate cut-off values for N (1.34), S (1.27) and M (1.36) as appropriate. Correlations between SARS-CoV-2 N, S and M antibody levels were calculated in GraphPad using Pearsons correlation coefficient. **(J)** Summary of R^2^ values for each correlation measured.

**Figure 5.**
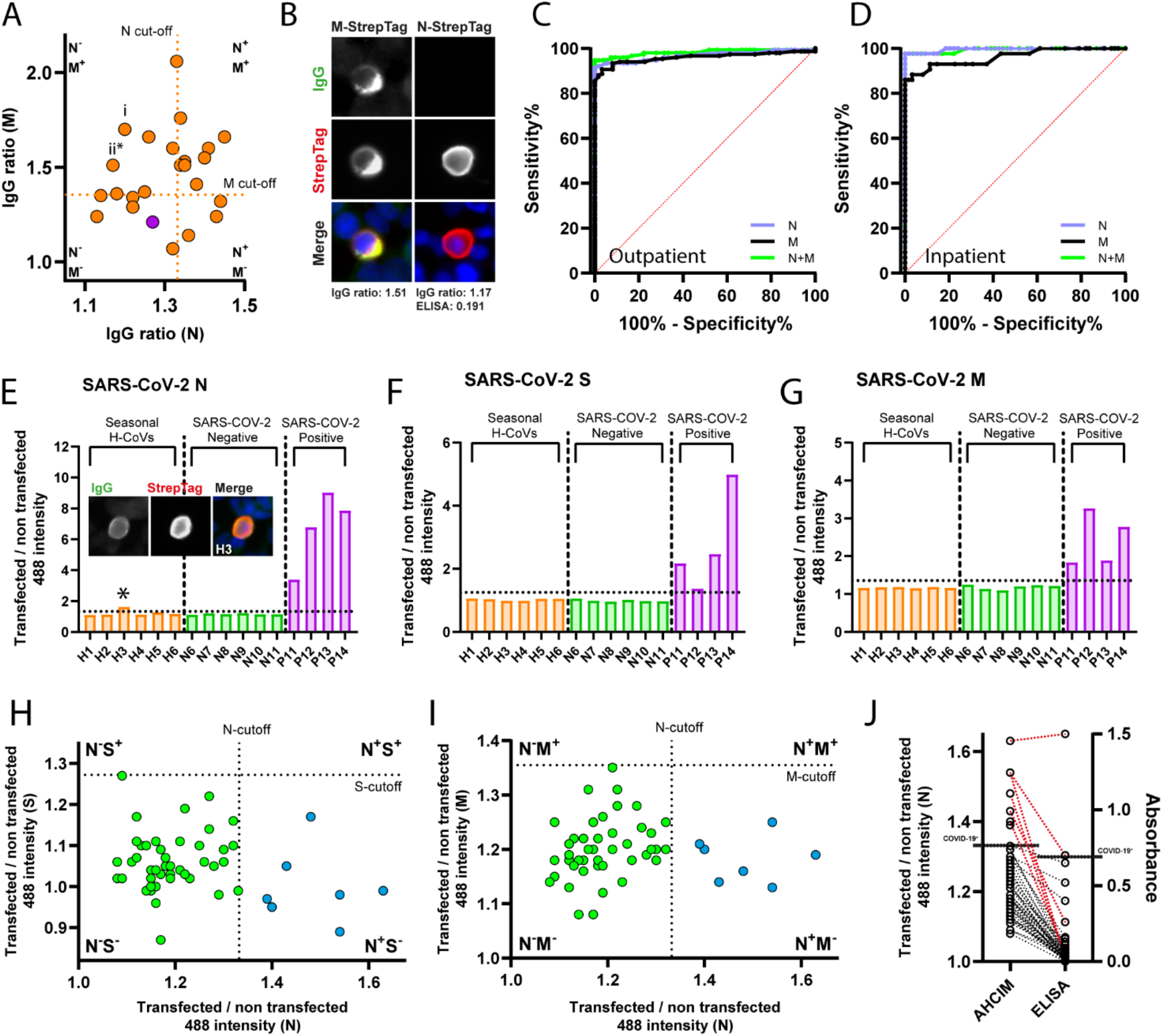
Improved SARS-CoV-2 serological sensitivity through detection of M IgG levels in subthreshold N samples. To determine if there were samples which were positive for M but lacked significant N reactivity (COVID-19^RT-PCR+^; M^AHCIM+^ N^AHCIM-^) we plotted M and N IgG ratios for the weakest samples **(A)**. Dashed orange lines indicating cut-off values for M and N respectively. COVID-19^RT-PCR+^;M^AHCIM+^ N^AHCIM-^ samples labelled (i) and (ii) fell below the ELISA threshold for classification as COVID-19 positive. **(B)** Representative images of sample (ii) assayed for IgG reactivity with M-StrepTag and N StrepTag. To determine if addition of M IgG ratios to N increased the sensitivity of AHCIM based detection of prior SARS-CoV-2 infection, N and M IgG ratios were summed together for each individual sample and optimal sensitivity and specificity values at a defined threshold calculated. ROC curves comparing sensitivity and specificity values for AHCIM based detection of N, M or N and M combined are shown for analysis of **(C)** outpatients and **(D)** inpatients. To investigate whether our platform is impacted by a cross-reactivity with seasonal coronaviruses a small number of samples (H1-H6) were analysed against StrepTagged SARS-CoV-2 nucleocapsid **(E)** spike **(F)** and M **(G)**. No cross reactivity with SARS-CoV-2 spike or M proteins were observed. However, one sample H3 (highlighted with an asterix) had IgG signal to N which was above the threshold. Inset in **(E)** shows representative images for this sample. To explore whether other pre-pandemic samples also showed a similar cross-reactivity we plotted the IgG ratios for N against S **(H)** and M **(I)**. Samples highlighted in blue represent sera that is positive for N but negative for either SARS-CoV-2 S or M suggesting cross-reactivity to a seasonal coronavirus. **(J)** To examine whether high ELISA values were also seen in the same pre-pandemic samples with seasonal coronavirus cross reactivity identified by AHCIM, IgG ratios and ELISA absorbances from these samples were plotted against one another. Dashed lines between data points link IgG ratios and ELISA absorbances from individual samples. Red dashed lines indicate samples with above threshold N signal identified by AHCIM. Half-length black dashed lines in the ELISA or AHCIM columns indicate the respective thresholds for classification of samples as COVID-19 positive or negative by ELISA or AHCIM.

In our earlier analysis, we found that 5.61% of COVID-19 samples were misclassified as negative by N ELISA, owing either to limitations in the sensitivity of the method, or the absence of antibodies against N in these samples. To see if detection of M could complement ELISA based testing for N, we first identified cases in our sample set which were positive by AHCIM for M IgG but negative for N IgG. 5 samples matched these criteria (**Figure 5A**). Two of these samples had below threshold ELISA absorbance values and could be reclassified as COVID-19 positive when using the IgG ratio calculated by AHCIM based detection of M antibodies. These samples had visibly detectable IgG responses against M but not N (**Figure 5B**). Combined detection of N antibodies by ELISA and M antibodies by AHCIM correctly called 187 out of 196 COVID-19 positive samples, increasing sensitivity to 95.4% over the use of ELISA based detection of N alone (**Supplementary Table 4**, ELISA N sensitivity; 94.4%, 185/196). Although the number of samples reclassified as COVID-19 positive based on the detection of M IgG represents only 1% of the total sample set analysed here, application of serological assays which screen for M antibodies could potentially result in a significant increase in the number of detectable COVID-19 cases at the level of mass testing across populations.

### Detecting M can help reduce the number of false positives detected by N based serological assays

While establishing our serological platform we noticed that in a few instances we observed a cross-reaction with N when using pre-pandemic sera or samples collected from individuals exposed to seasonal human coronaviruses (H-CoVs) (**Figure 5E**). Interestingly, each of these serum samples contained no detectable antibodies against either SARS-CoV-2 S or M (**Figure 5F-I**). Consistent with the results seen by AHCIM, 3 of the pre-pandemic N positive samples also registered high ELISA readings, with additional high value ELISA absorbance values for N from the pre-pandemic serum set also showing high AHCIM ratios (**Figure 5J**). As the negative pre-pandemic sample set was collected prior to the emergence of SARS-CoV-2, these results suggest that the signal we are observing is caused by the presence of antibodies to other coronaviruses. We are not the only group to have observed cross-reactivity of pre-pandemic samples with SARS-CoV-2 N; early studies on population level infection rates in Hong Kong reported a 1.5% false positive rate in pre-pandemic sera assayed for antibodies against SARS-COV-2 N (To, Cheng et al. 2020), whilst many commercial platforms see significant overlap in N signal between weak COVID-19 positive serum samples and pre-pandemic negative serum samples that show higher signal relative to the average (National 2020, Whitcombe, McGregor et al. 2021). Thus, it is likely that population based serological data based solely on N will over-estimate the number of individual infected with SARS-CoV-2.

### M antibodies may persist for longer than N antibodies

N antibody levels have been reported to decline more quickly over time in comparison to S antibody levels, raising doubts over the usefulness of N as a long-term indicator of SARS-CoV-2 infection (Long, Tang et al. 2020, Ripperger, Uhrlaub et al. 2020, Fenwick, Croxatto et al. 2021, Whitcombe, McGregor et al. 2021). To investigate whether M antibodies may persist for longer than N post infection, we examined N and M antibody levels over time by grouping samples based on the timepoint at which they were taken post PCR test and calculating an average IgG ratio per group. We focussed our analysis to COVID-19 outpatients samples as the inpatient sample values were more variable for both N and M (**Figure 6A-D**). As expected, we observed a time dependent increase in IgG levels for both N and M over the first 41 days (Li, Huang et al. 2020, Long, Liu et al. 2020). However, the levels of N start to decline and reach a level which is approximately 1.7-fold lower than at its peak. In contrast, M antibody levels remained relatively stable throughout the time course and showed only a very minor decrease in signal.

**Figure 6.**
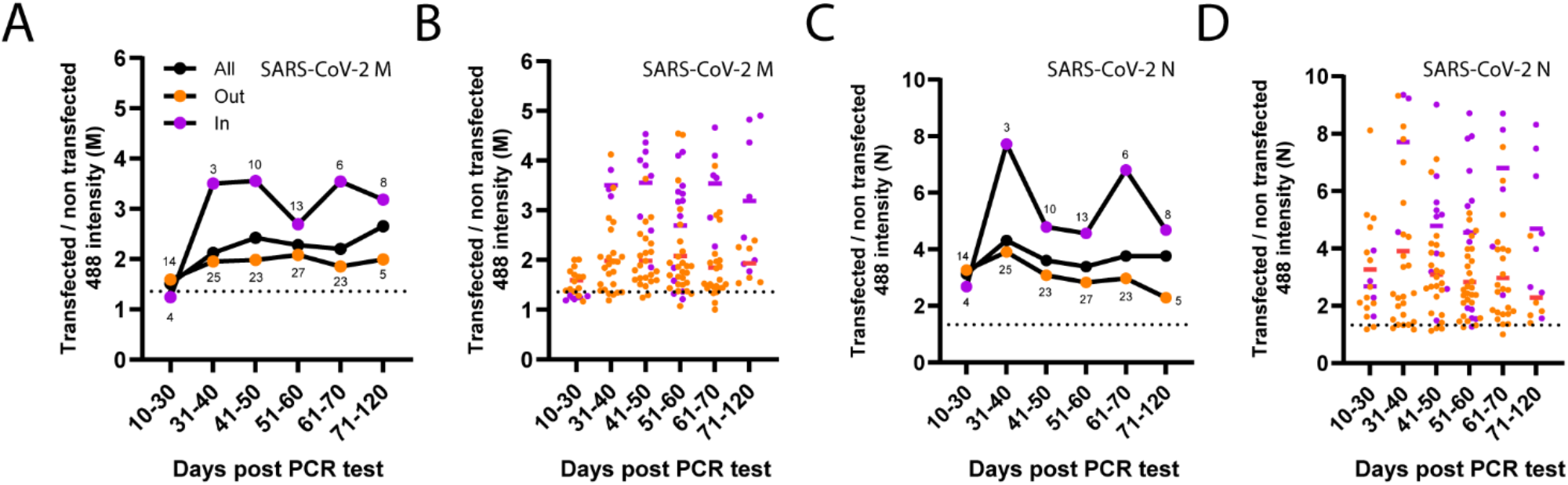
Analysis of M and N antibody levels over time. Samples were binned into groups spanning 10-day windows (except for samples obtained between days 10-30 or 71-120), depending on the time at which each serum sample was collected to determine M and N antibody concentrations over time. For each group, an average IgG ratio was calculated and plotted. Average ratios within each group are plotted for **(A)** M and **(C)** N. Individual samples within each group are plotted for M and N in **(B)** and **(D)** respectively. N numbers per group are plotted above inpatient and outpatient lines. Dashed lines Indicate cut-off values for classification of samples COVID-19 positive or COVID-19 negative.

## Discussion

The initial impetus for this paper was to investigate the utility of an immunofluorescence based serological screening platform for detection of N and S antibodies in sera from individuals infected with SARS-CoV-2. The method we used is routinely applied by countless laboratories worldwide for many purposes, involving transfection of plasmid encoded proteins into cells for transient expression, followed by a standard immunofluorescence staining protocol. Thus, if of acceptable sensitivity and specificity, we reasoned other laboratories could apply this methodology for a variety of purposes related to SARS-CoV-2 serology. A high throughput microscopy-based method has been described to assay total SARS-CoV-2 antibody levels (Pape, Remme et al. 2021), involving infection of cells with SARS-CoV-2 and detection of total bound human IgG following incubation with patient serum. However, this method did not allow antibody responses against individual SARS-CoV-2 proteins to be analysed, and to date no studies have extensively examined antibody responses in human serum samples against individual SARS-CoV-2 proteins using a high throughput immunofluorescence microscopy-based system.

In our study, we found the use of automated high content microscopy to be an efficient method of rapidly screening patient sera for antibodies against SARS-CoV-2 N, S and M. The method we employed here is semi-quantitative, with IgG ratios generated from mean fluorescence intensity values per transfected and non-transfected cell. Strong antibody responses were indicated by high IgG ratios and these values were frequently associated with strong absorbance readings obtained for the same samples by ELISA. Using these ratios, we assessed the sensitivity and specificity of AHCIM for detection of N and S IgG. Sensitivity and specificity values for detecting S IgG were equivalent to those seen for absorbances measured by ELISA, whilst our immunofluorescence-based platform had a slightly lower sensitivity than that seen by ELISA for detection of N IgG, likely related to higher background staining of transfected cells in permeabilised samples (**Supplementary Figure 4A)**. Detection of SARS-CoV-2 N and S antibodies by automated high content immunofluorescence microscopy may thus be a cheaper, faster, alternative to ELISA based methods in the absence of access to the purified proteins required for ELISA. The fast, high throughput, semi-quantitative method we present here is an upgrade on previous immunofluorescence based serological screening methods which have largely involved low throughput qualitative assessment of antibody titres (Peiris, Lai et al. 2003, Wu, Chiu et al. 2004, Wolfel, Corman et al. 2020).

Validation of our automated system through detection of N and S gave us the confidence to investigate antibody responses in infected samples against additional SARS-CoV-2 proteins. We chose to focus on the transmembrane protein M, one of four SARS-CoV-2 structural proteins, as studies conducted during the SARS-CoV pandemic identified detectable antibody responses against the SARS-CoV M protein (Wang, Wen et al. 2003, He, Zhou et al. 2005). We could clearly detect antibodies against SARS-CoV-2 M in a significant number of COVID-19 patients; 84.7% of COVID samples tested had detectable IgG against M (166 out of 196 PCR validated COVID-19 patients). Consistent with the results presented here, a recent study investigating serum reactivity to the structural proteins of SARS-CoV-2 by flow cytometry also found a high prevalence of M antibodies in COVID-19 positive patient sera, although only approximately 50% of COVID-19 patients tested contained M antibodies (Martin 2021).

The high number of COVID-19 patients with antibodies to M detected in this report and not in other studies (Jiang, Li et al. 2020, Shrock, Fujimura et al. 2020) is likely related to the state in which viral proteins are assayed for reactivity with patient sera. The M protein is a multi-spanning transmembrane protein proposed to be phosphorylated at its C terminus (Bouhaddou, Memon et al. 2020). The high amino acid sequence similarity (91% identity) to the membrane glycoprotein M of SARS-CoV suggests SARS-CoV-2 M also has the potential to be glycosylated (Oostra, de Haan et al. 2006). Both the folded architecture of M and the post translational modifications it is subjected to are likely to be key parts of epitopes recognised by circulating M antibodies. Underestimates of the seroprevalence of SARS-CoV-2 M antibodies from studies using viral peptide or proteins arrays are potentially related to the absence of post-translational modifications and folded epitopes in the versions of proteins and peptides used in these investigations (Hachim, Kavian et al. 2020, Jiang, Li et al. 2020, Shrock, Fujimura et al. 2020, Li, Xu et al. 2021). Thus, plasmid-based expression of M in mammalian cells holds several advantages in this respect as binding of antibodies to M is assayed with the protein in its native state. In addition, the StrepTagged-M used in isolation here is Golgi localised and therefore amenable to the same post translation modifications M undergoes in infected cells following its synthesis and transport through the secretory pathway (Oostra, de Haan et al. 2006, Bouhaddou, Memon et al. 2020).

The high seroprevalence of M antibodies in COVID-19 patients suggests that screening for M antibodies may be of great value to SARS-CoV-2 serological testing. The utility of M is underscored by complications in antibody testing arising from the widespread use of the S protein as the immunogen used in vaccination programs. Vaccinations combining S and N as immunogens to confer sterilizing immunity have also been proposed and tested (Dangi, Class et al. 2021). This strategy would prevent the use of both N and S antibody tests and thus further highlights the potential value of M as an additional serological marker. Currently however, detection of M antibodies would be most powerful when deployed alongside N to confirm prior SARS-CoV-2 infection in individuals vaccinated using S. In support of the use of M for this purpose, we identified cases from our sample set classified as COVID-19 negative based on the lack of detectable N antibodies which could be re-classified as positive through screening for M antibodies by AHCIM (**Figure 5A**). In addition, we also found that 11% of pre-pandemic COVID-19 negative samples contained antibodies that cross-reacted with SARS-CoV-2 N, but not S or M (**Figure 5H-I**). Detection of N antibodies in serum collected from both vaccinated and un-vaccinated patients may therefore not indicate prior infection with SARS-CoV-2 in 100% of cases. With these caveats in mind, complementation of ELISA based detection of N antibodies with detection of M antibodies by AHCIM could provide an immediate strategy to enhance the sensitivity and specificity of SARS-CoV-2 serological testing during the era of mass COVID-19 vaccinations. Future work to identify the key immunodominant epitopes of M detected by AHCIM may simplify serological testing using M even further, allowing the production of a SARS-CoV-2 M protein compatible for use with an ELISA format, thereby helping to facilitate widespread serological testing using M.

Another key issue in SARS-CoV-2 serology relates to the length of time that each serological marker persists for post infection (Meyer 2021). Long lived serological markers are desirable for optimal COVID-19 sero-surveillance as the loss of detectable antibody levels after infection may lead to an underestimation of infection rates and indicate a loss of protective immunity. Whilst detection of sero-conversion due to SARS-CoV-2 infection in those vaccinated is reliant on assaying for N antibodies, the utility of N as a long-lived serological marker is uncertain (Long, Tang et al. 2020, Ripperger, Uhrlaub et al. 2020, Fenwick, Croxatto et al. 2021, Whitcombe, McGregor et al. 2021), although recent data suggests that N antibodies levels remain high for longer than previously believed (Shrotri, Harris et al. 2021). Elucidating the kinetics of M antibody levels over time may help to establish whether M may be a more durable serological marker of SARS-CoV-2 infection than N. A preliminary analysis of N and M IgG ratios plotted against days post PCR test when each serum sample was collected showed a trend in N, but not M, for antibody levels to decline over time, however the number of samples used in this analysis at crucial later timepoints (71-120 days) was low (**Figure 6**). While this data is suggestive of greater temporal stability in M antibody levels, longitudinal profiling of N and M antibody titres in individual COVID-19 serum samples would likely give a clearer answer as to the utility of M as a long-lived serological marker in comparison to N.

Our observations regarding the seroprevalence of M IgG also raise several other important questions which warrant further investigation. Correlations between the presence of M antibodies and disease outcomes and whether antibodies against M may serve to provide and bolster the neutralising activity of humoral immune responses generated against SARS-CoV-2 are currently unknown and are clearly important questions to address. Of note, M IgG ratios were significantly higher in inpatient samples analysed in this study (**Figure 3J**) suggesting higher M antibody levels correlate with more severe COVID-19 cases, as previously reported for N and S (Roltgen, Powell et al. 2020, Batra, Tian et al. 2021, Yates, Ehrbar et al. 2021) and seen again here in this study (**Figure 3B & F**).

The synergistic use of proteome-wide tagging of viral proteins with automated high content immunofluorescence microscopy could provide a rapid standardised format to quickly identify the most antigenic proteins of newly emergent pathogens. This would be of value for both vaccine development and guiding which antigens to use as markers of prior infection in diagnostic methods such as ELISA or AHCIM. One could in theory quickly progress from obtaining the complete genome of a previously uncharacterised pathogen, to generating a codon optimised tagged library of proteins for expression in mammalian cells, to identifying the strongest antigenic targets across the whole proteome using an automated immunofluorescence microscopy-based platform like the one described here. The identification of M as a key target of the humoral immune response to SARS-CoV-2 provides a first-hand example of the advantages of such an approach.

## Data Availability

All data produced in the present study are available upon reasonable request to the authors.

## Acknowledgements

This work was funded by the generous support of the Sheffield Teaching Hospitals NHS Foundation Trust and an unrestricted charitable donation from The Danson Foundation (DAN581929). A. Peden, D. Williams and A. Shun-Shion are supported by the BBSRC (BB/S009566/1 and BB/J014443/1). T. de Silvia was supported by Wellcome Trust Intermediate Clinical Fellowship (110058/Z/15/Z). J. Edgar is supported by a Sir Henry Dale Fellowship jointly funded by the Wellcome Trust and the Royal Society (216370/Z/19/Z). We are grateful to Professor David James (University of Sheffield) for production of the purified mammalian spike protein used for ELISA. We are also grateful to Dr Xiaoming Fang (University of Sheffield) for useful comments and advice on the manuscript.

## Supplementary figures

**Supplementary Figure 1.**
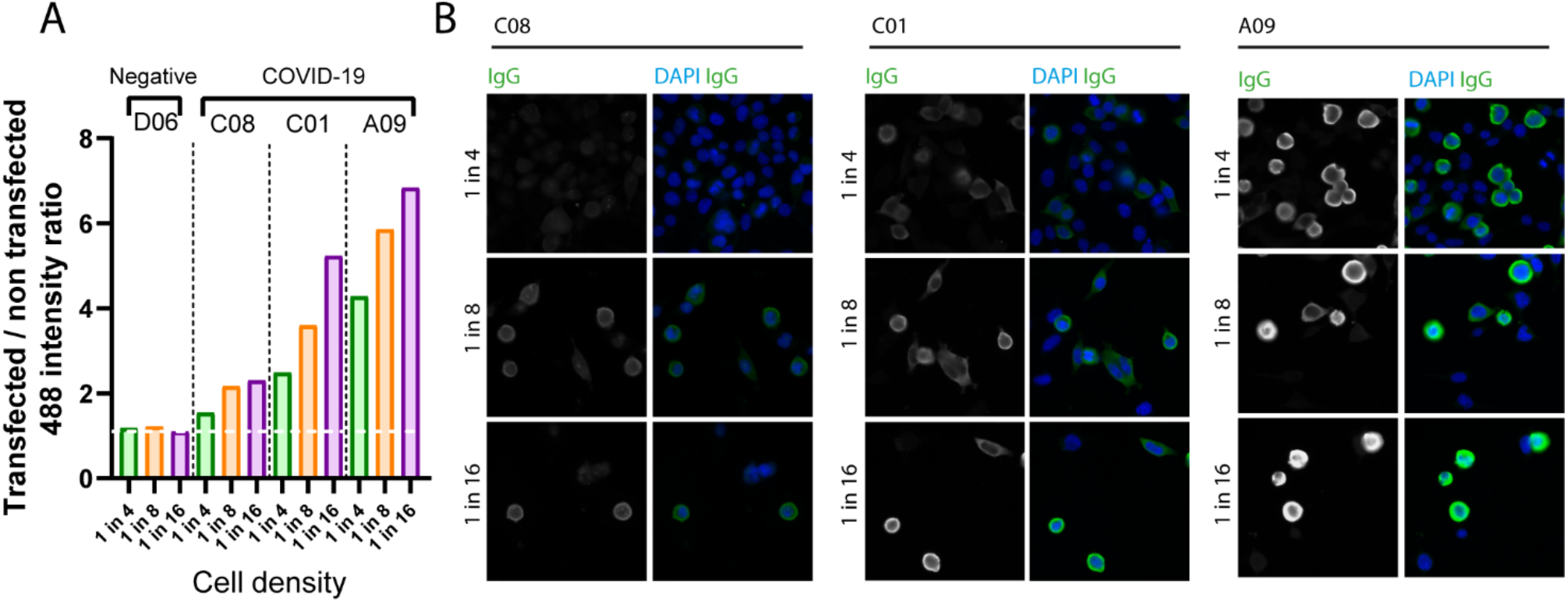
Decreasing the number of transfected cells improves 488-intensity ratios. **(A)** Representative images of StrepTagged SARS-CoV-2 nucleocapsid transfected cells imaged at different ratios of transfected to non-transfected cells. Serum samples with strong (A09), intermediate (C01) and weak (C08) SARS-CoV-2 nucleocapsid IgG responses were selected. Transfected cells plated at 100% confluency were split at the indicated dilutions, fixed, incubated with patient sera and processed for immunofluorescence as described. (**B)** Automated quantification of 488 intensity ratios for COVID19 serum samples A09, C01 and C08 and the COVID19 negative serum sample D06. Decreasing cell density increases the IgG ratio for strong, intermediate and weak COVID-19 positive samples.

**Supplementary Figure 2.**
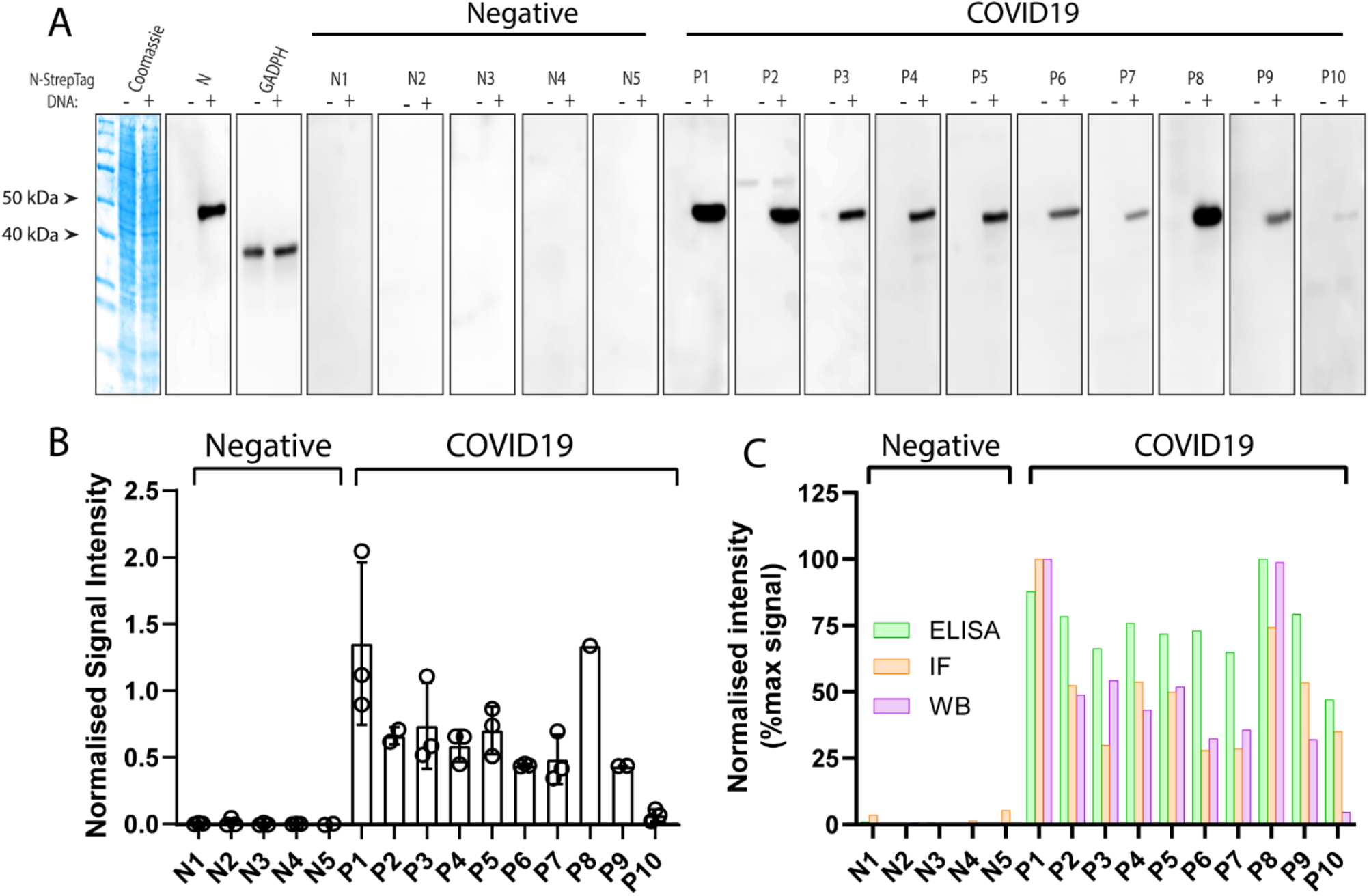
Detection of SARS-CoV-2 nucleocapsid IgG by western blot. **(A)** HEK293 cell pellets transfected with either StrepTagged SARS-CoV-2 nucleocapsid (+) or empty vector (-) for 48 hours were lysed directly into SDS-PAGE sample buffer, lysates resolved by SDS-PAGE and transferred to PVDF. Individual strips of membrane were probed with sera from the training set 15 and bound human IgG detected with HRP-labelled anti-human IgG secondary antibodies. **(B)** Quantification of signal detected by western blotting for each of the training set 15 serum samples. N=3 for each sample, unless otherwise indicated by *. Not all serum samples were able to be analysed 3 times due to sample availability. (**C)** Comparison of normalised signal strength for each serum sample from the training set 15 as measured by either western blotting, immunofluorescence or ELISA.

**Supplementary Figure 3.**
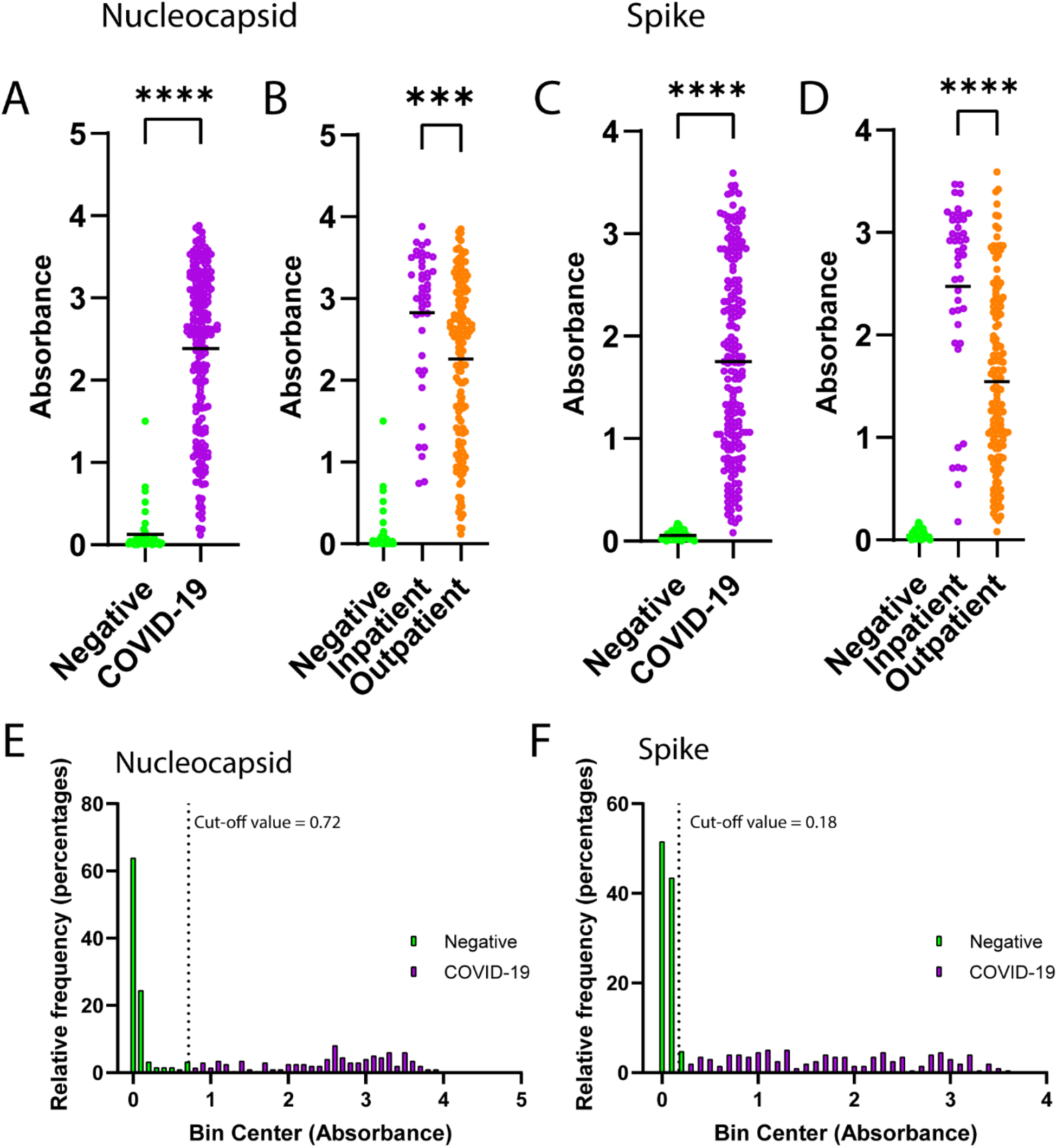
ELISA readings performed on same serum sample set analysed by AHCIM. Individual ELISA absorbance values for COVID-19 negative (n=62) and COVID-19 (n=196) positive serum samples measured against purified nucleocapsid **(A)** or purified spike **(C)**. Stratification of serum samples based on inpatient (n=44) and outpatient (n=152) status for nucleocapsid **(B)** and spike **(D)**. Inpatient antibody levels are also higher than outpatient IgG levels for nucleocapsid and spike, as seen for AHCIM. Frequency distribution histograms of ELISA absorbance values from pre-pandemic and COVID-19 positive serum samples for **(E)** nucleocapsid and **(F)** spike. Samples were binned into groups of 0.1 absorbance value units width and plotted. Dashed lines indicate positive and negative cut-off value calculated from generated ELISA ROC curves. \*\*\*\**P*<0.0001, \*\*\**P*<0.001 (unpaired t test).

**Supplementary Figure 4.**
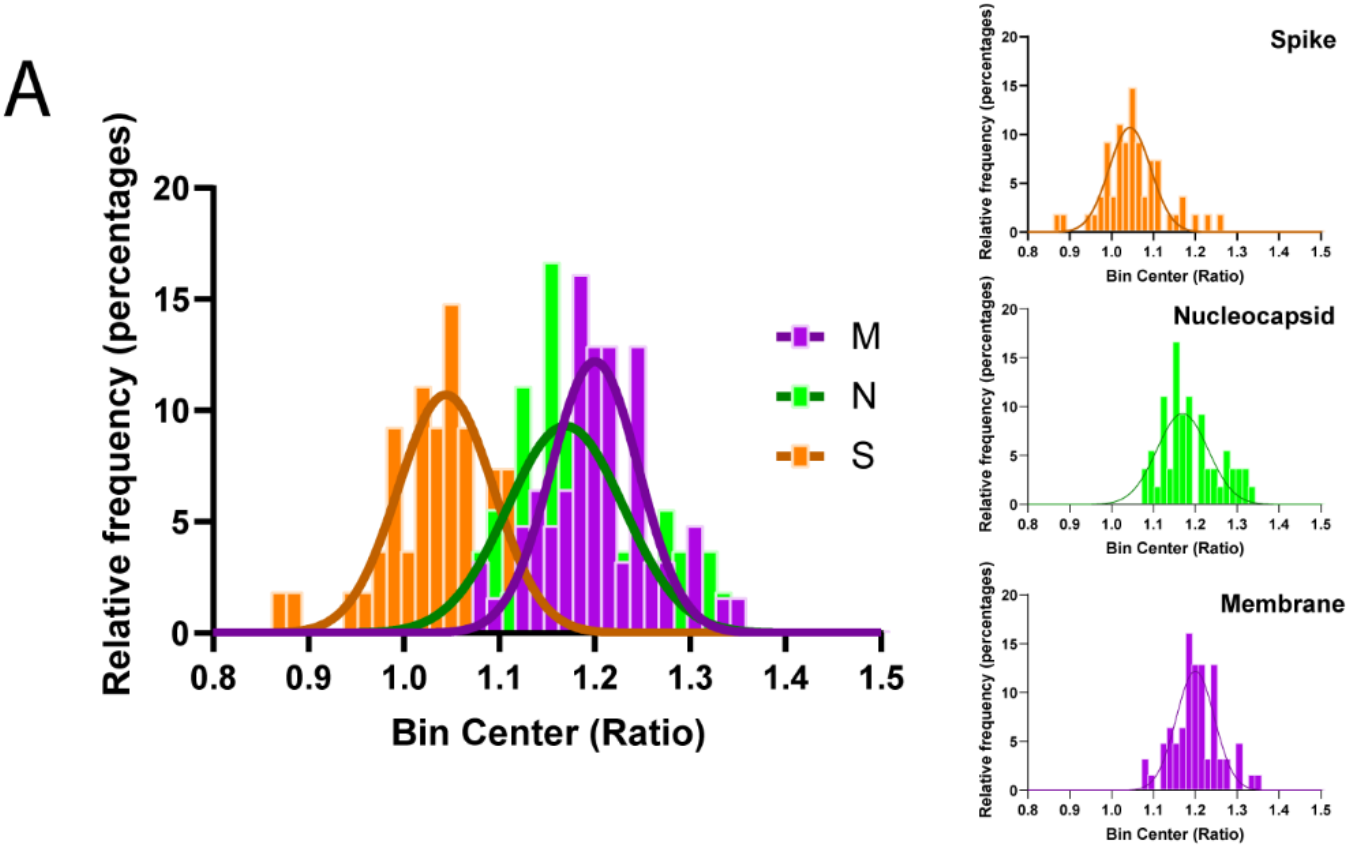
Comparison of the distribution of IgG ratios for spike, nucleocapsid and M COVID-19 negative samples. **(A)** Frequency distribution histograms for negative samples IgG ratios from spike, nucleocapsid and M. Curves for nucleocapsid and M are shifted to the right indicating higher background fluorescence in transfected cells. Bin width was set at 0.15 ratio points.

**Supplementary Figure 5.**
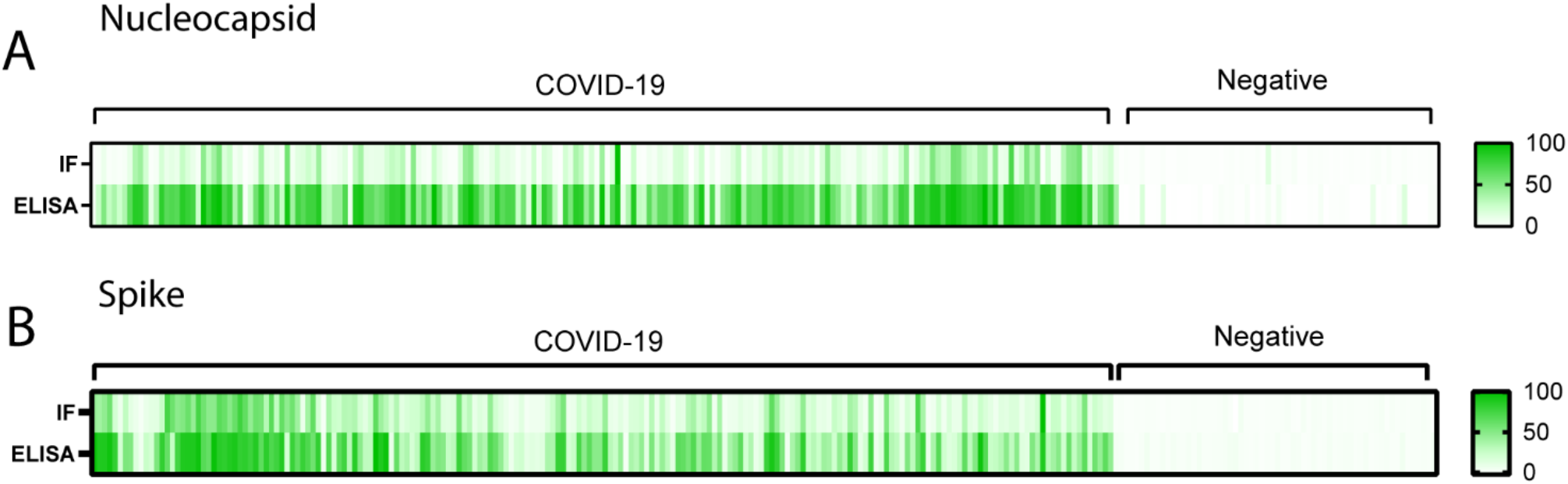
Normalised AHCIM IgG ratios and ELISA absorbance values from individual serum samples show a high degree of concordance. Heatmap comparing normalised ELISA and AHCIM values for individual pre-pandemic and COVID-19 positive serum samples from **(A)** nucleocapsid and **(B)** spike data sets. Data sets were normalised by setting the highest measured value from respective AHCIM and ELISA measurements as 100% and expressing all other samples as a percentage of maximum signal relative to this value.

**Supplementary Figure 6.**
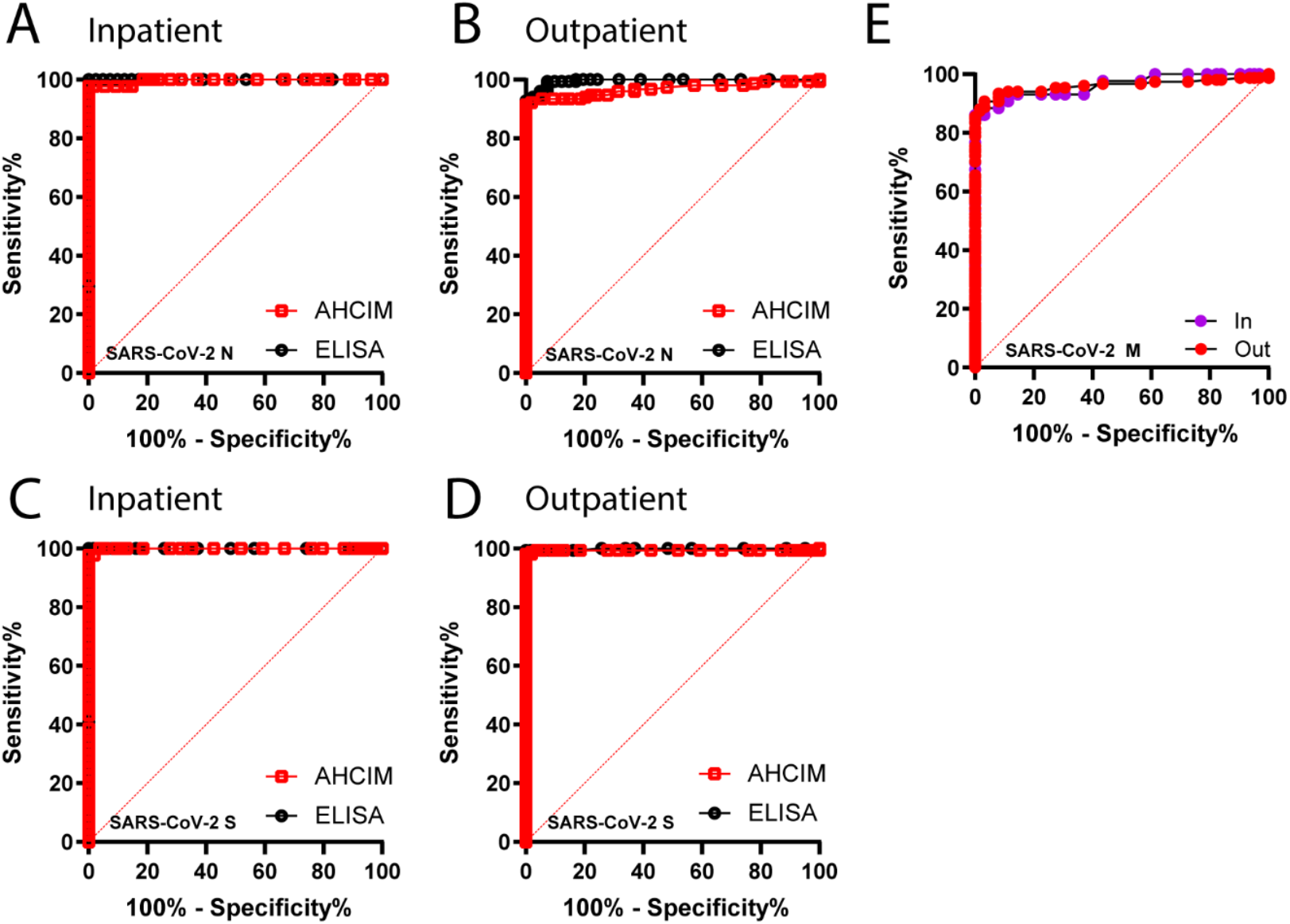
Performance of ELISA and AHCIM based spike and nucleocapsid IgG detection in discriminating between negative and COVID-19 serum samples. ROC curves for nucleocapsid and spike were generated using the 62 negative and 196 positive samples described in Figure 3. ROC curves comparing the performance of ELISA and AHCIM nucleocapsid and spike IgG detection for stratified inpatient **(A & C)** and outpatient **(B & D)** data from the same larger sample set. **(E)** ROC curve analysis of M IgG ratios for inpatient and outpatient samples.

**Supplementary Table 1.**
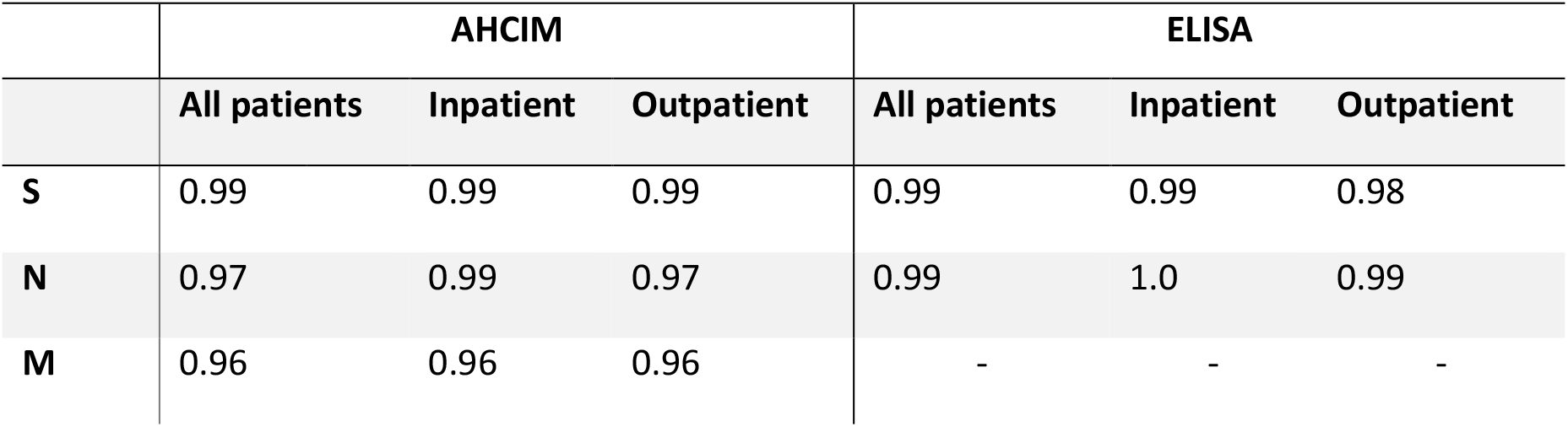
Summary of ROC curve AUC values generated by ELISA or AHCIM for N and M.

|  | AHCIM |  |  | ELISA |  |  |
| --- | --- | --- | --- | --- | --- | --- |
|  | All patients | Inpatient | Outpatient | All patients | Inpatient | Outpatient |
| **S** | 0.99 | 0.99 | 0.99 | 0.99 | 0.99 | 0.98 |
| **N** | 0.97 | 0.99 | 0.97 | 0.99 | 1.0 | 0.99 |
| **M** | 0.96 | 0.96 | 0.96 | - | - | - |

**Supplementary Table 2.**
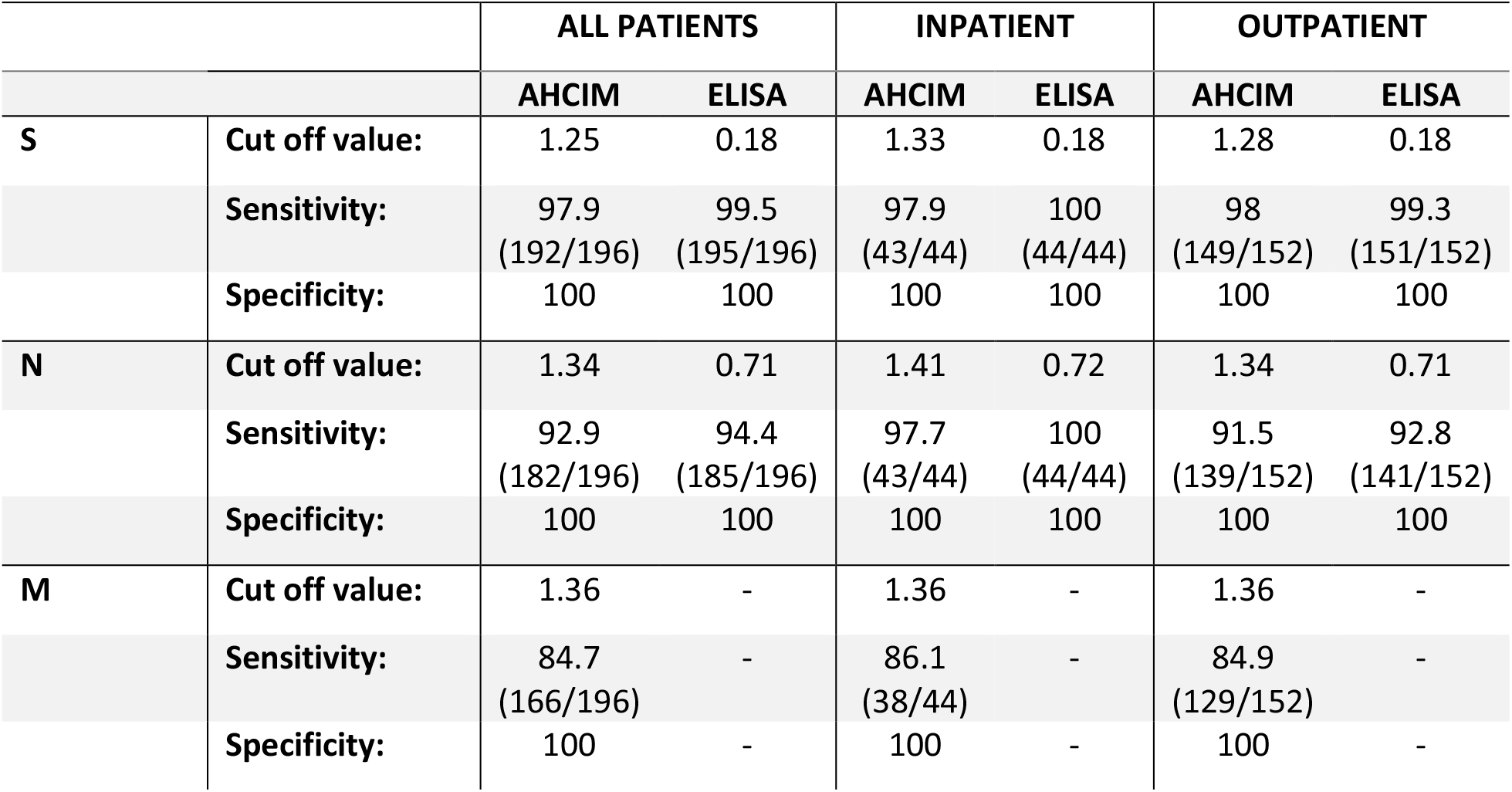
Summary of sensitivity and specificity values for N S and M based on cut-off values generated from AHCIM and ELISA ROC curves.

**Supplementary Table 3.** Sensitivity and specificity values for AHCIM based detection of M and N individually, or M and N combined.

|  |  | AHCIM CUT-OFF<br>VALUE | SENSITIVITY | SPECIFICITY | AUC |
| --- | --- | --- | --- | --- | --- |
| ALL | N | 1.34 | 92.9<br>(182/196) | 100 | 0.97 |
|  | M | 1.35 | 84.7<br>(166/196) | 100 | 0.96 |
|  | N + M | 2.61 | 95.4<br>(187/196) | 100 | 0.98 |
| IN | N | 1.41 | 97.7<br>(43/44) | 100 | 0.99 |
|  | M | 1.36 | 86.1<br>(38/44) | 100 | 0.96 |
|  | N + M | 2.73 | 97.4<br>(43/44) | 100 | 0.99 |
| OUT | N | 1.34 | 91.5<br>(139/152) | 100 | 0.97 |
|  | M | 1.36 | 84.9<br>(129/152) | 100 | 0.96 |
|  | N + M | 2.61 | 94.7<br>(144/152) | 100 | 0.98 |

**Supplementary Table 4.** Sensitivity and specificity values based on combined detection of N antibodies by ELISA and detection of M antibodies by AHCIM.

|  |  | SENSITIVITY | SPECIFICITY | CUT-OFF VALUES |
| --- | --- | --- | --- | --- |
| ALL | N | 94.4<br>(185/196) | 100 | 0.71 ELISA |
|  | N + M | 95.4<br>(187/196) | 100 | 0.71 ELISA, 1.36 AHCIM |
| IN | N | 100<br>(44/44) | 100 | 0.72 ELISA |
|  | N + M | 100<br>(44/44) | 100 | 0.72 ELISA, 1.36 AHCIM |
| OUT | N | 92.8<br>(141/152) | 100 | 0.71 ELISA |
|  | N + M | 94.1<br>(143/152) | 100 | 0.71 ELISA, 1.36 AHCIM |

